# Frailty progression following severe infections in adults ≥65 years in US and England: two matched cohort studies

**DOI:** 10.64898/2026.03.13.26348319

**Authors:** Kwabena Asare, Kathryn E. Mansfield, Georgia R. Gore-Langton, Sharon L. Cadogan, Eleanor Barry, Ruth Keogh, Vincent Lo Re, Maria C. Rodriguez-Barradas, Amy C. Justice, Christopher T. Rentsch, Charlotte Warren-Gash

## Abstract

**Background:** We investigated frailty progression after severe infections in adults (≥65 years) in the US and England.

**Methods:** We conducted parallel matched cohort studies using: US Veterans Aging Cohort Study (VACS-National, 2008-2019; median age 74 years; 98% male); and English Clinical Practice Research Datalink (2006-2019; median age 76 years; 45% male). Adults hospitalised primarily for infection (i.e., severe infection) were matched in calendar date order to individuals without severe infection on age, sex, care site, and US only, plus race and ethnicity. We measured frailty using VACS Index 2·0 (US) and Electronic Frailty Index (eFI; England). We estimated annual conditional mean frailty differences between adults with versus without severe infection using linear regression adjusting for baseline frailty, demographics, lifestyle factors, infection history, and US only, comorbidities.

**Results:** Mean baseline frailty was higher in those with severe infection than those without (US: 57 v 48; England: 0·17 v 0·12). At Year 1, adjusted mean frailty was higher among adults with severe infections than those without (US: VACS Index +2·0, 95% CI 1·9-2·0; England: eFI +0·005, 95% CI 0·005-0·006). At Years 2-5, adjusted mean frailty remained higher after severe infection; however, compared to Year 1, differences were smaller in US, and larger in England. Effects varied by infection type (strongest for lower respiratory tract infections, meningoencephalitis (UK only), urinary tract infections, and sepsis).

**Interpretation:** Individuals with severe infections had higher frailty at baseline and follow up than those without. Preventing both frailty and infections is important for improving health in older age.

**Funding:** Wellcome

**Research in context:** *Evidence before this study:* We searched PubMed (inception to October 27, 2025), for published articles evaluating the association between infections and frailty, with no language restrictions. We used the search terms [(infection OR infectious) AND (frailty OR frail)]. We found fifteen observational studies investigating associations between individual infections (including: HIV, cytomegalovirus, SARS-CoV-2, acute respiratory infection, urinary tract infection, and influenza) and frailty in adults. Frailty measures varied: eight studies used Fried’s phenotype index, six used versions of the cumulative deficit index (i.e., Edmonton Frail Scale, FRAIL-NH Scale, Hospital Frailty Risk Score, Clinical Frailty Score, Veterans Affairs Frailty Index, Vulnerable Elders Survey-13), and one study used the Timed Up and Go Test. Results from identified studies were mixed, with nearly half (7/15) reporting a positive association between the infection studied and frailty, and the remaining eight finding no evidence of association. In cross-sectional analyses, HIV, SARS-CoV-2, cytomegalovirus, and urinary tract infection, were each associated with higher mean frailty scores or frailty prevalence. In longitudinal analysis, hospitalisation for acute respiratory infection was followed by higher mean hospital frailty risk scores two years post-discharge. SARS-CoV-2 infection was associated with early onset (i.e., higher hazard) of frailty over three years follow-up. However, other studies found no association between HIV, SARS-CoV-2, acute respiratory infection and influenza, and frailty prevalence, incidence, or transition between frailty states. These mixed findings may reflect methodological differences between the studies, including variation in frailty measures, and study limitations. Frailty exists along a continuum of vulnerability, and progression after infection may be an important outcome, yet current evidence is scarce. It remains unclear whether severe infections or different types of infection, are associated with faster frailty deterioration. Similarly, it is uncertain whether post-infection frailty risk varies by pathogen (bacterial, viral, parasitic, fungal), infection type (sepsis, urinary tract infection, skin and soft tissue infection, meningitis/encephalitis, lower respiratory tract, gastroenteritis), or by age, sex, social deprivation, and pre-existing comorbidities.

*Added value of this study:* Our study compared frailty progression over a five-year period between adults aged ≥65 years with severe infection (hospitalisation primarily due to infection) versus comparators without severe infection. We found higher baseline frailty at severe infection onset than in matched comparators. We saw evidence of increased frailty progression over time in people following severe infections compared to those without, however, these differences were small. We also saw higher risk of worsening frailty progression in older adults and those with dementia. Further, worsening frailty progression varied by infection type (strongest for lower respiratory tract infections, meningoencephalitis (UK only), urinary tract infections, and sepsis).

*Implications of all the available evidence:* Our findings underscore the importance of both frailty and infection prevention in improving health in older age. Additional studies are required to explore other wider life-course influences on frailty, to guide the development of comprehensive preventive strategies.

## Introduction

Frailty describes overall resilience and capacity to recover quickly following health problems.^1^ Frailty typically affects older people due to age-related decline, however up to a third of adults over 85 years do not experience frailty.^1^ Disparities in frailty development and progression can manifest in adults of the same age raising questions about its underlying causes and mechanisms.^1,2^ As the global population ages, understanding factors contributing to frailty progression is important to inform healthy ageing interventions.^1^

Infections, particularly severe infections, may trigger health status decline, with consequent frailty exacerbation and reduced short-to-medium-term physical function,^1,3–5^ (although existing studies are conflicting).^6–8^ Functional decline after infections may also accelerate dementia onset.^1,9,10^ However, it is unclear whether infections or specific infection types influence longer-term frailty trajectories, particularly as frailty predisposes to subsequent infections and poor infection outcomes.^11–14^

Therefore, we explored whether frailty worsens over a five-year period among individuals following severe infections (i.e., hospitalised primarily due to infection) after age 65 years compared to matched comparators with no severe infection in the US and England.

## Methods

### Study design, population, and data sources

We used routinely collected electronic health records (EHR) from adults aged 65 years and above in the US and England for two matched cohort studies.

In the US, we used the Veterans Aging Cohort Study (VACS-National) (01-01-2008 to 31-12-2019). VACS-National includes >14.1 million individuals (95% male reflecting service personnel demographics) who ever received care in the Department of Veterans Affairs (VA).^15,16^ The VA is the largest integrated healthcare system in the US serving >9 million Veterans annually at >1300 hospitals, medical centres, and community outpatient clinics.^15,16^ As part of their health monitoring, US Veterans are expected to have annual clinical check-ups, which typically include routine laboratory tests.

In England, we used primary care data from practices contributing to the Clinical Practice Research Datalink (CPRD) Aurum (March 2025 build) (01-04-2006 to 31-12-2019). Aurum includes pseudonymised data covering nearly 25% of UK population, and is broadly representative of the UK population on age, sex, ethnicity, and region.^17,18^ We also used linked: 1) hospital admissions data from Hospital Episode Statistics – Admitted Patient Care (HES-APC) to identify individuals with severe infection;^19^ and 2) Index of Multiple Deprivation (IMD) data.^17^

In both studies, participants were eligible for inclusion from the latest of: study start (US, 01-01-2008; England, 01-04-2006); 65th birthday; or one year after registration with health care site (to reliably establish baseline health status). We randomly matched without replacement, in calendar time order, individuals with any severe infection (i.e., first hospital admission primarily due to infection after age 65) with up to five individuals with no such record after age 65 (i.e., without severe infection) on age, sex, health care site (i.e., England primary care practice, US VA site of care), and, in the US only, race and ethnicity. Follow-up continued until the earliest of: 1) study end (31-12-2019); 2) death; 3) last health care visit (US); 4) last data collection from practice (England); or 4) for matched comparators only, their first severe infection (when they entered as exposed participants).

### Exposure

We defined severe infection as the first infection that was the primary reason for hospitalisation after age 65 years. The hospital admission date was used as the index date. Unexposed participants with no record of hospitalisation primarily due to infection after age 65 years were assigned the same index date as their matched infection-exposed counterparts.

In the US, we defined severe infection as the first principal inpatient diagnosis for infection in a hospital admission after age 65 years coded with relevant International Classification of Diseases, Tenth Revision, Clinical Modification (ICD-10-CM) or ICD-9-CM codes. In England, we used the first hospital admission after age 65 years with an ICD-10-coded infection diagnosis recorded in the first diagnostic position of any episode of the admission (for administrative reasons, England hospital admissions are divided into one or more episodes under the care of a specific hospital consultant).

### Outcomes

Our outcome was frailty, defined using different validated measures in the US and England (**Text S1**). In the US, we used VACS Index 2.0. The VACS Index is a physiologically-based index originally developed in people living with HIV and validated for predicting all-cause mortality in people living with and without HIV.^20^ VACS Index scores typically range between 0-100, with higher scores associated with higher mortality risk.^20,21^ A five point increment in VACS Index 2.0 is associated with a 6·5% increased 5-year mortality risk.^21^ VACS Index 2.0 is calculated based on weights (from five-year-all-cause-mortality Cox regression model) for the following covariates: age, body mass index (BMI), and individual laboratory test results (i.e., albumin, haemoglobin, white blood cell count, liver fibrosis 4, alanine aminotransferase, platelet count, estimated glomerular filtration rate, CD4, HIV-1 RNA, hepatitis C infection).^20,21^ To calculate VACS Index at index date, we looked backward for the most recent records from 2 years up to 14 days before index date (to avoid the infection influencing baseline VACS Index). In each subsequent year of follow-up, we calculated VACS Index based on the most recent recorded measures within the past 12 months.

In England, we measured frailty using Version one of the Electronic Frailty Index (eFI).^22^ EFI was developed and validated in England for predicting all-cause mortality, hospitalisation, and nursing home admission.^22^ It uses a cumulative model of equally weighted primary care morbidity coding across 36 deficits. Baseline eFI was defined using deficits recorded (based on codes or prescriptions), at any time prior to the 14 days before index date. We assessed frailty at each follow-up year (1-5), updating deficit status annually. A deficit was recorded as present, and eFI updated, if it appeared within the previous 12 months; otherwise, the previous year’s status was carried forward.

### Covariates

We used directed acyclic graphs to help identify potential confounders of the relationship between severe infection and subsequent worsening frailty (**Figure S1**). All covariates were based on records on or before index date (**Text S2**). We identified four broad categories of potential confounders: demographics, socio-economic disadvantage, lifestyle, and underlying health. In each setting, our modelling strategy was influenced by data availability and missingness, and relationship with frailty index.

In the US, we included the following potential confounders: 1) demographics: age, sex, race and ethnicity, VA site of care (matching factors); 2) socio-economic disadvantage: rural-urban area of residence, Area Deprivation Index^23^; 3) lifestyle: alcohol use and use disorder, smoking; 4) underlying health: outpatient-diagnosed infection (in 5 years before cohort entry), Charlson Comorbidity Index (CCI).

In England, we considered the following potential confounders: 1) demographics: age, sex, practice site (matching factors); 2) socio-economic disadvantage: individual-level quintiles of IMD (where missing, supplemented with practice-level IMD)^17^; 2) lifestyle: harmful alcohol use, smoking; and 3) underlying health: primary-care recorded infection in the five years before index date. CCI was not included for England because frailty index (eFI) calculation includes comorbidities. We only adjusted for ethnicity in sensitivity analyses due to relatively high proportion (i.e., 40%) of missingness.

We considered age, sex (England only), care home residency (England only), deprivation, dementia, diabetes mellitus, pathogen and infection type, and CCI (US only) as potential effect modifiers. We categorised infections by pathogen (i.e., bacterial, viral, fungal, parasitic) and infection type (i.e., sepsis, urinary tract infection (UTI), skin and soft tissue infection (SSTI), meningoencephalitis, lower respiratory tract infection (LRTI), gastroenteritis) based on ICD codes available online (<u>infection codes and classifications</u>).

### Statistical analysis

We initially summarised descriptive characteristics at index date, follow-up time, and frailty index at follow-up by severe-infection exposure status.

We used separate linear regression models to estimate conditional mean difference in frailty at each year of follow-up (i.e., Year 1, 2, 3, 4, and 5) between adults with and without severe infection. We accounted for matching with cluster-robust (by matching-id) standard errors using Generalised Estimating Equations with independent correlation structure. We adjusted each regression model for baseline frailty and then sequentially adjusted for confounders in the following order: 1) demographics, 2) socioeconomic disadvantage, 3) lifestyle, and 4) underlying health.

#### Sensitivity analyses

We explored how robust our results were by repeating our main analyses in a series of sensitivity analyses (**Table S1**).

#### Secondary analyses

In separate analyses in both the US and England, we stratified by the following potential effect modifiers assessed at cohort entry: age group, sex, quintiles of Area Deprivation Index (US), quintiles of IMD (England), care home residency (England), dementia, diabetes mellitus, CCI categories (US), infection pathogen and infection type. We compared sub-group effects for evidence of statistical interaction with linear hypothesis tests.

Analytical R code^24^ and code lists used to define study variables are available online (<u>codelists</u>, analytical R code: <u>US</u>, <u>England</u>). Both analyses were approved by the London School of Hygiene & Tropical Medicine Research Ethics Committee (References: US, 31029; England, 31298). Additionally, the US study was approved by the institutional review boards of Yale University (Reference: #1506016006) and VA Connecticut Healthcare System (Reference: #AJ0013), and the English study by CPRD’s Independent Scientific Advisory Committee (Reference: 24_004305).

### Role of the funding source

The study funders had no role in study design, data request and access, analysis, interpretation, or manuscript writing.

## Results

### US

In the main analysis we included 1,066,018 adults who had VACS Index recorded at index date and at least once during follow-up (**Figure 1**). These were 229,311 with severe infection and 836,707 matched counterparts without severe infection. The distribution of matching factors (age, sex, race and ethnicity), and some other characteristics (i.e., rural or urban area of residence, deprivation) were similar between groups (**Table 1**). Alcohol use and use disorder was lower among individuals with severe infection compared to those without (moderate-to-high-risk consumption 9·2% v. 12·2%). Both comorbidity burden and smoking were higher among adults with severe infection than those without (current smoker: 28% v. 21%; CCI 3+: 45% vs. 17%) (**Table 1**). VACS Index at index date was nine points higher among adults with severe infection (57, standard deviation [SD] 14) than those without (48, SD 11).

**Figure 1.**
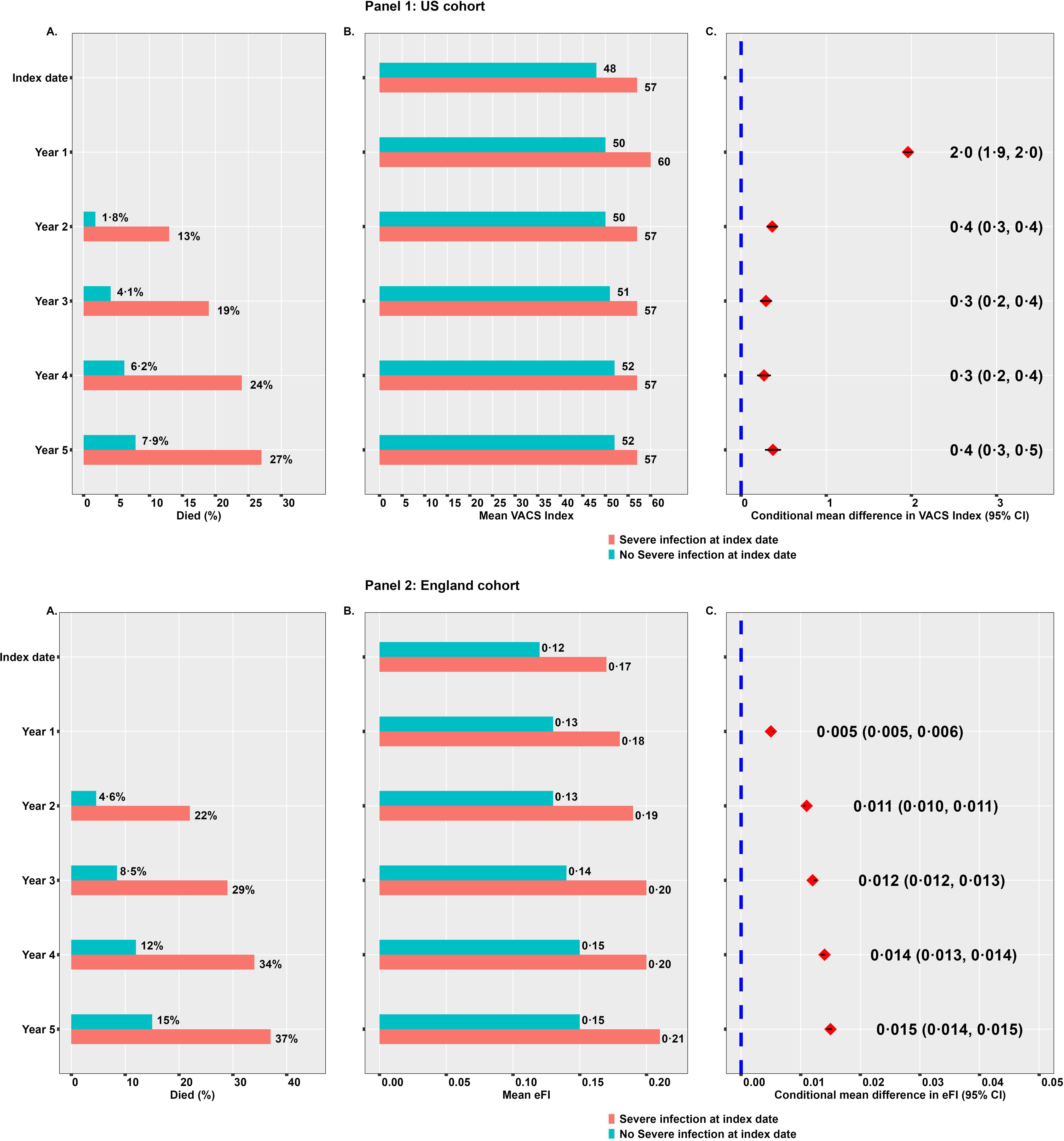
Flow diagram of the study cohorts in the US and England. * Participants were eligible for inclusion from the latest of: study start (US, 01-01-2008; England, 01-04-2006); 65th birthday; or one year after registration with general practitioner’s practice or VA care site (to reliably establish baseline health status). LRTI=Lower respiratory tract infection, SSTI=Skin and soft tissue infection, UTI=Urinary tract infection, VA = Department of Veterans’ Affairs, VACS = Veterans Ageing Cohort Study.

**Table 1.**
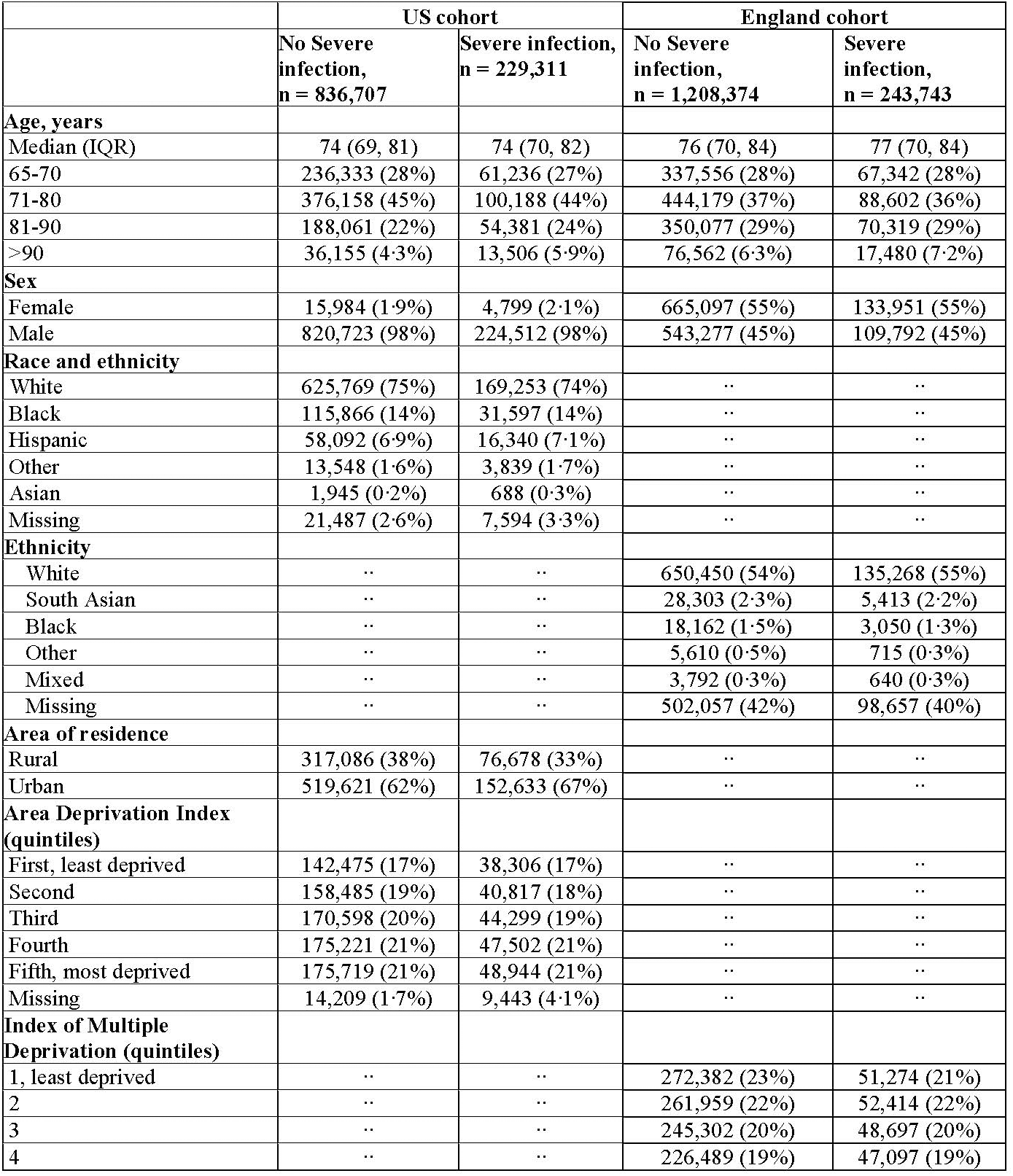
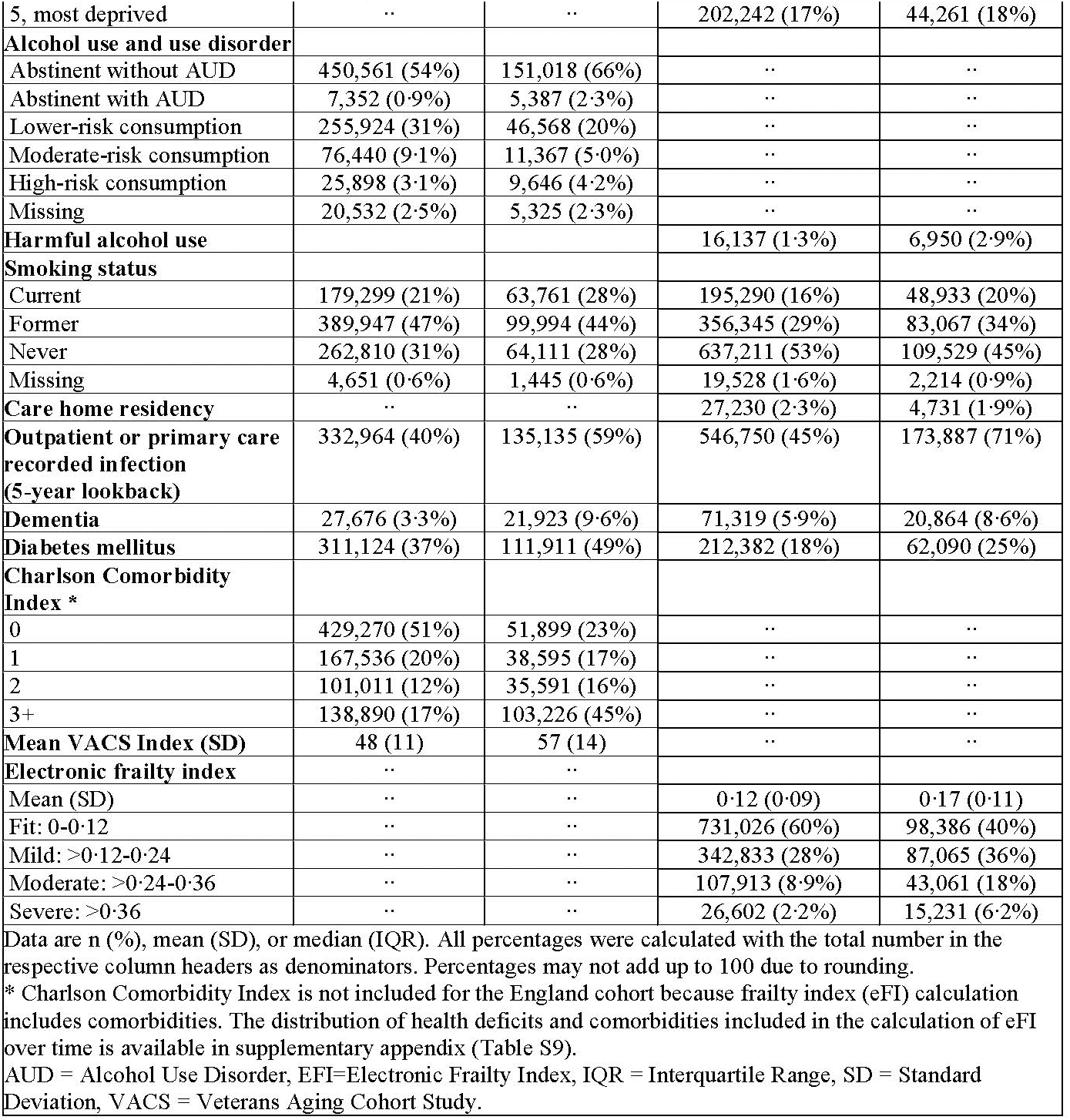
Characteristics of adults at index date.

Adults with severe infection had shorter follow-up (median 2·7 years, interquartile range [IQR] 1·0, 5·2) than counterparts without severe infection (median 3·8 years, IQR 2·0, 6·4) (**Table 2**). By design, almost all adults, with and without severe infection were available at first year follow-up. However, from follow-up Years 2 to 5, a higher proportion of adults with severe infection were censored largely because they had died (Year 2: 13% with infection died vs 1·8% without, Year 3: 19% vs 4·1%, Year 4: 24% vs 6·2%, Year 5: 27% vs 7·9%) **(Figure 2**).

**Figure 2.**
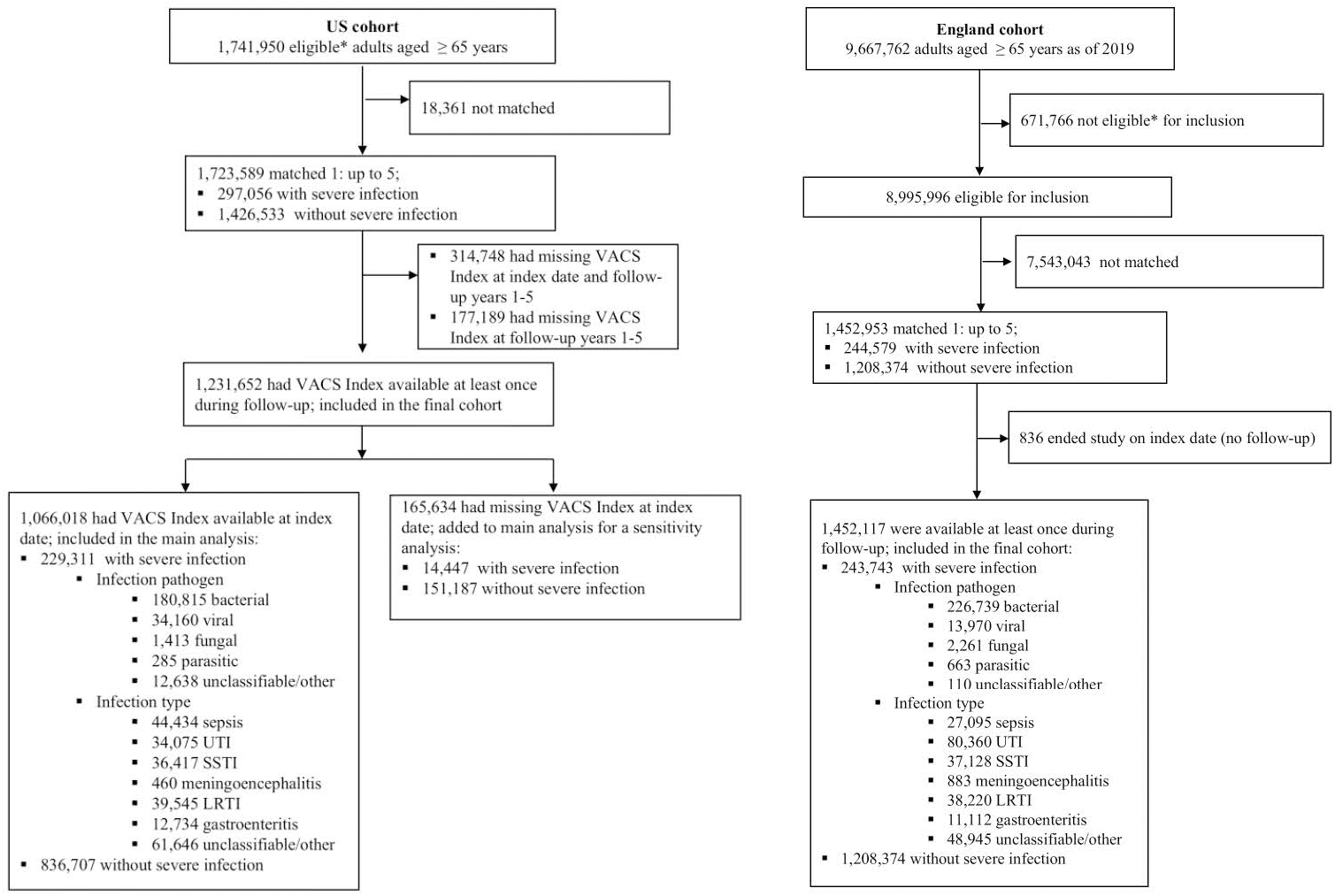
Plots showing results from index date to follow-up years 1 to 5 in adults aged 65 years and above with and without severe infection. **Panel 1:** US cohort. A. Censoring due to death, B. Mean VACS Index, and C. Conditional mean difference in VACS Index (95% CI), between adults with versus without severe infection at index date. The VACS Index typically ranges from 0 to 100. The conditional mean difference in VACS Index is adjusted for VACS Index at index date and other variables at index date; age, sex, race and ethnicity, rural or urban residence, quintiles of Area Deprivation Index, alcohol use and use disorder, smoking status, outpatient recorded infection (5-year lookback), and Charlson Comorbidity Index. **Panel 2:** England cohort. A. Censoring due to death, B. Mean Electronic Frailty Index (eFI), and C. Conditional mean difference in eFI (95% CI), between adults with versus without severe infection at index date. The eFI is a proportion of 36 deficits and hence ranges from 0 to 1. The conditional mean difference in eFI is adjusted for eFI at index date and other variables at index date; age, sex, quintiles of multiple deprivation index, harmful alcohol use, smoking status, and primary care recorded infection (5-year look back).

Conditional mean differences in frailty attenuated with each sequential confounder adjustment (**Table S3)**. After full adjustment, estimated conditional mean differences in VACS Index between adults with severe infection compared to those without were: Year 1: 2·0 (95% CI 1·9, 2·0), Year 2: 0·4 (95% CI 0·3, 0·4), Year 3: 0·3 (95% CI 0·2, 0·4), Year 4: 0·3 (95% CI 0·2, 0·4), and Year 5: 0·4 (95% CI 0·3, 0·5) (**Figure 2**). All mean differences at follow-up were greater than zero, indicating a higher mean VACS Index at follow-up in adults with severe infection than those without.

### England

We included 1,452,117 adults aged 65+ years: 243,743 with severe infection and 1,208,374 matched counterparts without severe infection (**Figure 1**). The distribution of age and sex (i.e., matching factors), ethnicity, and level of deprivation were similar among adults with and without a severe infection (**Table 1**). Harmful alcohol use and smoking were higher among adults with severe infection than those without (harmful alcohol use: 2·9% v. 1·3%; smoking: current smoker 20% v. 16%). Mean eFI at index date was 0·5 points higher among adults with severe infection (0·17, SD 0·11) than counterparts without (0·12, SD 0·09).

Adults with severe infection contributed less follow-up (median 2·1 years, IQR 0·5, 5·0) than counterparts without severe infection (median 4·0, IQR 1·8, 7·2) (**Table 3)**. All adults were available at one year follow-up. But, from Years 2 to 5 a high proportion of adults with severe infection were censored because they had died (Year 2: 22% with infection died vs 4·6% without, Year 3: 29% vs 8·5%, Year 4: 34% vs 12%, Year 5: 37% vs 15%) (**Figure 2)**.

Conditional mean differences were stable when we sequentially adjusted for potential confounders (**Table S4)**. Comparing those with and without severe infections, fully adjusted conditional mean differences in eFI (95% CI) during follow-up were: Year 1: 0·005 (0·005, 0·006), Year 2: 0·011 (0·010, 0·011), Year 3: 0·012 (0·012, 0·013), Year 4: 0·014 (0·013, 0·014), and Year 5: 0·015 (0·014, 0·015) (**Figure 2)**.

### Sensitivity analyses

Results of sensitivity analyses were consistent with those of the main analyses (**Tables S3, and S4)**. In the US, characteristics of the cohort including those with missing VACS index at index date (used in sensitivity analysis exploring impact of their exclusion) were broadly similar to the main analysis cohort with VACS Index available at index (**Tables S5, and S6**).

### Secondary analyses

In the US, conditional mean differences in VACS Index (95% CI) during follow-up Years 1-5 in adults with severe infection compared to those without, was higher among sub-groups of older adults (Year 1: **65-70 years**: 1·3 [1·2, 1·4]; **71-80 years**: 1·8 [1·7, 1·9]; **81-90 years**: 2·7 [2·6, 2·8]; **>90 years**: 3·8 [3·5, 4·0]), and higher in adults with dementia (Year, 1: 3·1 [2·9, 3·3]) than those without (Year 1: 1·8 [1·8, 1·9]) (**Table S7)**. We also found evidence that conditional mean differences in VACS Index (95% CI) varied by the exposed individual’s infection type; Year 1: **LRTI**: 2·9 (2·7, 3·0); **UTI**: 2·6 (2·4, 2·7); **sepsis**: 3·0 (2·9, 3·1); **Gastroenteritis**: 2·0 (1·9, 2·0); **SSTI**: 1·7 (1·6, 1·8); **Meningoencephalitis**: 0·4 (−0·8, 1·5). We found no evidence of effect modification by deprivation, diabetes, CCI, and infection pathogen.

In England, the conditional mean differences in eFI (95% CI) during follow-up Years 1-5 varied by infection type of the exposed individual; Year 1: **LRTI**: 0·007 (0·007, 0·008)**; UTI**: 0·007 (0·006, 0·007); **Meningoencephalitis**: 0·006 (0·004, 0·008); **SSTI**: 0·005 (0·005, 0·005); **Gastroenteritis**: 0·004 (0·003, 0·004)**; sepsis**: 0·003 (0·003, 0·004); and lower among adults with dementia (Year 1: 0·002 [0·001, 0·002]) than those without dementia (Year 1: 0·006 [0·006, 0·006]) (**Table S8**). Effects were also lower in adults who were residing in care homes (Year 1: 0·001 [0·000, 0·002]) than those not in care homes (Year 1: 0·005 [0·005, 0·006]). We found no evidence of effect modification by age, sex, deprivation, diabetes, or infection pathogen.

## Discussion

Our findings from parallel studies in two national health systems show higher frailty at baseline and during each of five years follow-up in adults hospitalised primarily due to infection after age 65 years (severe infection) compared to matched individuals without severe infection. In the US, the difference in frailty was highest in the first year of follow-up, with effect estimates attenuating from Year 2. In England, the difference in frailty was smallest during the first year of follow-up and increased progressively from Year 2. The association between severe infection and higher frailty was modified by infection type (strongest for LRTI, followed by meningoencephalitis, UTI, and sepsis), dementia and, in the US only, age.

Our findings of higher mean frailty at baseline among adults with severe infection are consistent with existing evidence from cross-sectional studies. A US cross-sectional analysis without confounder control in adults aged 65+ years reported higher mean frailty scores among participants with recurrent UTI than those without.^25^ Other cross-sectional studies have demonstrated associations between SARS-CoV-2^4^, cytomegalovirus^26^ and HIV^3,27^ infections, and higher frailty prevalence. Our baseline findings may represent bidirectional associations between severe infection and frailty development, consistent with other studies.^28–31^

There are few longitudinal studies investigating the association between specific infections and change in frailty. In one cohort study, individuals aged 80+ years hospitalised for acute respiratory infection had a larger mean increase in frailty two years after discharge than comparators aged 80+ years hospitalised for cataract surgery.^5^ The study reported no estimates comparing mean difference in frailty change between the exposure groups.^5^ In another longitudinal study SARS-CoV-2 infection was associated with earlier onset (i.e., higher hazard) of categorical frailty during three years of follow-up.^32^ Higher prevalence of frailty and pre-frailty have also been observed after hospitalisation with SARS-CoV-2 infection.^33^

To our knowledge, ours is the first longitudinal study to compare difference in frailty during follow-up between adults with and without severe infection. Our study used large routinely collected datasets from two national health systems with different approaches to funding healthcare (including England, where the study population was representative of England population). By addressing the same question in two different data sources with different strengths and limitations we were able to triangulate our findings.^34^ We included all diagnosed infections instead of single infections recorded in the electronic health records of older adults from both settings and explored effect modification by infection pathogen and infection type. We adjusted for a broad range of potential confounders including baseline frailty. Our findings were robust in sensitivity analyses.

In defining severe infection exposure, we only included infections recorded as the principal/primary diagnosis during hospitalisation which may have misclassified individuals some individuals as comparators who actually had severe infection potentially leading to underestimation of difference in frailty. Further, our severe infection-exposure definition will have misclassified participants with community-acquired or managed infections not leading to hospital admission as unexposed. However, we aimed to address the relationship between severe infection and frailty progression. Infections not leading to hospital admission, were likely to be mild, so any impact is likely to mean our results were underestimates. Likewise, our findings were consistent in sensitivity analyses where we included only adults with evidence of community-acquired infections and their matched comparators.

CPRD is representative by age, sex, deprivation and geographical spread, hence selection bias in England may not be a major concern.^17^ Conversely, individuals in the VA tend to be older, have more chronic health conditions and risk behaviours compared to the US general population, and are predominantly male, potentially limiting generalisability of our results to the wider US population. But evidence indicates overall disease burden does not differ between Veterans and non-Veterans after adjustment for key demographic and geographic factors included in the analysis.^35^ Our findings are also consistent with prior studies in non-VA populations, suggesting limited concerns regarding generalisability.^5,32^ Another key source of selection bias was censoring over time largely driven by death and higher among individuals with severe infection. The increased death in those with severe infections could have underestimated the difference in frailty over time, as comparisons were largely among survivors of severe infection (who were more likely to be healthy).

In England, using eFI to measure frailty made it impossible to capture frailty improvement as eFI Version 1 only ever increases over time.^22^ It is also likely that participants with severe infection had more healthcare contacts and were therefore more likely to have deficits recorded. Our results may therefore overestimate the effect of infection on frailty, due to differential ascertainment of frailty status in people with and without infection. In the US, using the VACS Index allowed us to capture dynamic changes in frailty over time. All individuals in the VA are expected to have annual clinical check-ups for health monitoring and hence less likely to have differential VACS Index ascertainment. Nevertheless, the choice of baseline frailty ascertainment window of up to 14 days prior to infection recording may have captured early infection-related changes in some individuals.

We saw a lower prevalence of dementia in the US, where its effect on frailty after infection was detrimental. Differences in baseline prevalence across the two settings may reflect heterogeneity in the likelihood of dementia diagnosis and recording.^36^ The unexpected finding that frailty progressed more after infections in individuals without dementia in England may reflect differences in frailty ascertainment: there may be limited clinical benefit to recording accumulating frailty deficits among individuals with dementia in England, whereas physiologic frailty is recorded routinely each year in the US.

We are unable to exclude residual confounding as a potential explanation for our findings. There are some potential confounders that we would be unable to capture robustly using routinely collected data. For example, clinicians do not routinely record social engagement, carer support or access to occupational or physical therapies, which may all play an important role in the relationship between severe infections and frailty progression.

Our results suggest that frailty may already be present at the onset of severe infections in older age and it is likely to worsen slightly over time among individuals who experience severe infections compared to those who do not. The adverse impact of severe infections on frailty progression was greatest following LRTI, meningoencephalitis (UK only), UTI, and sepsis. In the US only, we saw stronger effects among older adults and those with dementia. These groups may therefore represent priority targets for preventative and post-infection interventions aimed at mitigating worsening frailty progression. Infection prevention with vaccination as well as early detection and prompt management could help alleviate frailty deterioration in old age. Further research is needed on broader life-course determinants of frailty to inform holistic measures for frailty prevention.

### Contributors

KA, KEM, CTR, and CWG conceptualised and designed the study. KA, KEM, RK, EB, CTR, and CWG developed the statistical analysis plan. KA and CTR extracted and verified the data. KA, KEM, GRGL, SLC, EB, RK, CTR, and CWG had access to the data through their affiliation with London School of Hygiene and Tropical Medicine. KA managed, cleaned, and analysed the data. KEM, CTR, and CWG reviewed the statistical analysis. All authors interpreted the data. KA, KEM, and CWG wrote the initial draft. All authors reviewed and approved the manuscript. CWG is the guarantor. All authors are responsible for the decision to submit for publication.

### Declaration of interests

All authors declare no competing interest.

### Data sharing

We do not have the rights to publicly share the data. However, Due to US Department of Veterans Affairs (VA) regulations and our ethics agreements, the analytic data sets used for this study are not permitted to leave the VA firewall without a data use agreement. This limitation is consistent with other studies based on VA data. However, VA data are made freely available to researchers with an approved VA study protocol. For more information, please visit https://www.virec.research.va.gov or contact the VA Information Resource Center at. Data used for England cohort are available on request from CPRD. The code used to clean, and analyse the data are publicly available on the GitHub repositories (<u>US</u>, <u>England</u>)

## Data Availability

We do not have the rights to publicly share the data. However, Due to US Department of Veterans Affairs (VA) regulations and our ethics agreements, the analytic data sets used for this study are not permitted to leave the VA firewall without a data use agreement. This limitation is consistent with other studies based on VA data. However, VA data are made freely available to researchers with an approved VA study protocol. For more information, please visit https://www.virec.research.va.gov or contact the VA Information Resource Center at. Data used for England cohort are available on request from CPRD. The code used to clean, and analyse the data are publicly available on the GitHub repositories (US, England)

## Acknowledgements

This study is based in part on data from the Clinical Practice Research Datalink obtained under licence from the UK Medicines and Healthcare products Regulatory Agency. The data is provided by patients and collected by the NHS as part of their care and support.

Funding for this study is through a Wellcome Career Development Award (225868/Z/22/Z) received by CWG. The Veterans Aging Cohort Study is supported by the National Institute on Alcohol Abuse and Alcoholism [P01-AA029545, U01-AA026224, U24-AA020794, U01-AA020790, U10-AA013566].

---

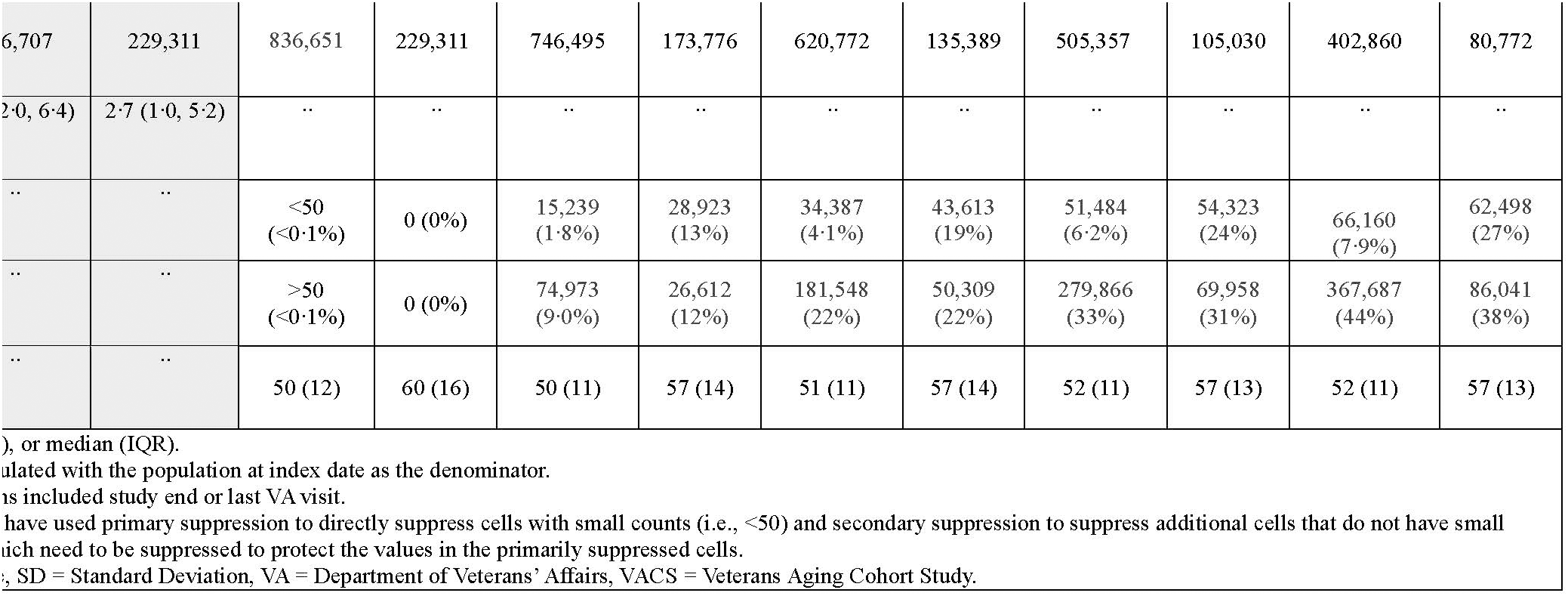

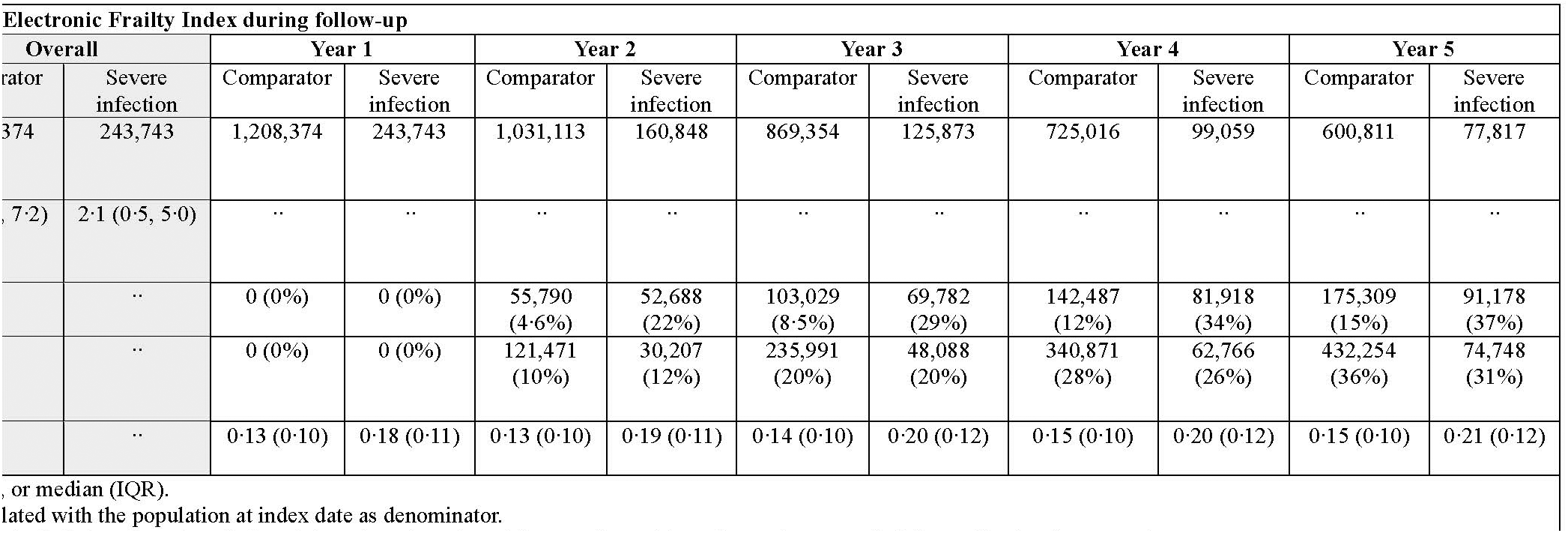

## Notes

### Competing Interest Statement

The authors have declared no competing interest.

### Author Declarations

Both analyses were approved by the London School of Hygiene & Tropical Medicine Research Ethics Committee (References: US, 31029; England, 31298). Additionally, the US study was approved by the institutional review boards of Yale University (Reference: #1506016006) and VA Connecticut Healthcare System (Reference: #AJ0013), and the English study by CPRD Independent Scientific Advisory Committee (Reference: 24_004305).

