## Supplementary material for "Frailty progression following severe infections in adults ≥65 years in US and England: two matched cohort studies"

**Supplementary Appendix for Frailty progression following severe infections in adults ≥ 65 years in the US and England: two matched cohort studies**

**Supplementary Appendix 1: Supplementary Figures**

Figure S1 Directed acyclic graphic depicting potential confounders of the association between severe infections and frailty progression**.**

**Supplementary Appendix 2: Additional information for variable definitions**

Text S1 Outcomes

Text S2 Covariates

**Supplementary Appendix 3: Supplementary Tables**

Table S1 Description of sensitivity analyses conducted

Table S2 Supplementary characteristics of adults at index date

Table S3 US cohort: Linear regression for conditional mean differences in VACS Index at follow-up years 1 to 5 between adults with and without severe infection at index date

Table S4 England cohort: Linear regression for conditional mean differences in Electronic Frailty Index at follow-up years 1 to 5 between adults with and without severe infection at index date

Table S5 US cohort: Cohort characteristics at index date including adults with missing VACS Index

Table S6 US cohort: VACS Index during follow-up in cohort including adults with missing VACS Index at index date

Table S7 US cohort: Stratified linear regression for conditional mean differences in VACS Index at follow-up years 1 to 5 between adults with and without severe infection at index date

Table S8 England cohort: Stratified linear regression for conditional mean differences in Electronic Frailty Index at follow-up years 1 to 5 between adults with and without severe infection at index date

Table S9 England cohort: Prevalence of Electronic Frailty Index deficits over time

**Supplementary Appendix 1: Supplementary figures**

**Figure S1. Directed acyclic graphic depicting potential confounders of the association between severe infections and frailty progression·**

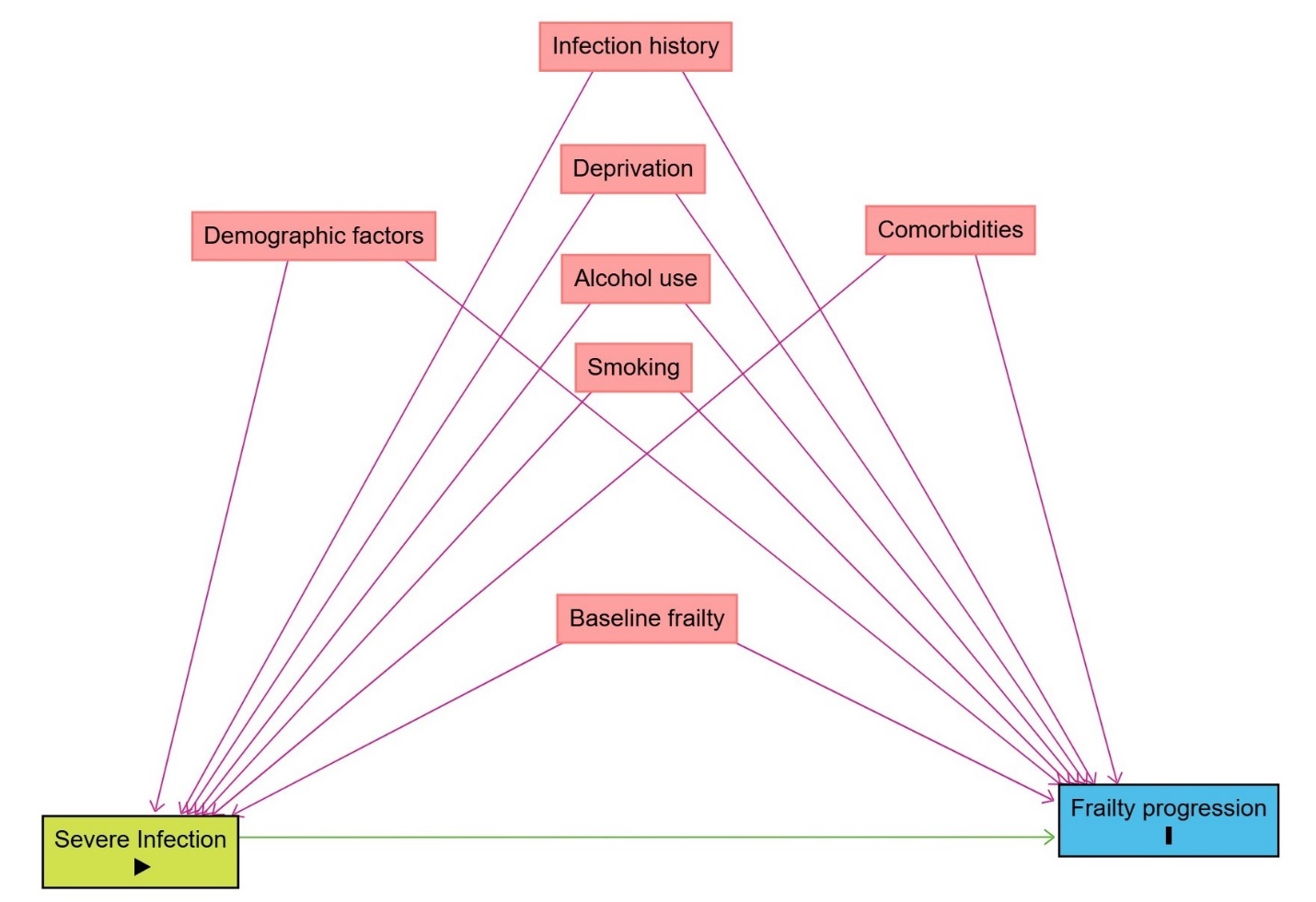

**Supplementary Appendix 2: Additional information for variable definitions**

**Text S1: Outcomes**

US cohort: VACS Index

Liver fibrosis 4 was calculated using aspartate transferase [AST], alanine aminotransferase [ALT], and platelet count. We set the following measures to population-expected reference values if only one of them was missing: albumin=4 g/dL, ALT=25 U/L, AST=25 g/dL, haemoglobin=14 g/dL, platelet=200 k/mL, white blood cell count=5.5 k/mL, and BMI=25kg/m^2^. If more than one of the measures was missing, we recorded VACS Index as missing. As most adults (~99%) were HIV negative, we set CD4 and log HIV-1 RNA to expected population reference values (CD4: 1000 cells/mm^3^, HIV-1: 1.3 log copies/mL) for all adults.

England cohort: Electronic Frailty Index

The Electronic Frailty Index (eFI) is calculated as the proportion of deficits an individual observes out of the full list of 36. We defined the polypharmacy deficit as having ≥5 prescribed medications, using British National Formulary Chapters 1-15.^1^ We defined the remaining thirty-five deficits using primary care morbidity coding (coded using Systematized Nomenclature of Medicine-Clinical Terms, SNOMED-CT, codes).

**Text S2: Covariates**

US cohort

Area Deprivation Index is a measure of US neighborhood disadvantage estimated quarterly based on education, employment, housing-quality, and poverty measures.^2^

We classified individuals into five mutually exclusive groups of alcohol use and use disorder based on recent Alcohol Use Disorder Identification Test Consumption (AUDIT-C) score, and ICD-9- or ICD-10-coded diagnoses of alcohol use disorder (AUD) within two years before index date.^3^

Smoking status (i.e., never, former, and current) was based on the most frequent response in the five years before index date. All individual comorbidities used to calculate CCI (i.e., Myocardial infarction, Congestive heart failure, Peripheral vascular disease, Cerebrovascular disease, Dementia, Chronic Obstructive Pulmonary Disease, Rheumatologic disease, Peptic ulcer disease, Mild liver disease, Diabetes mellitus , Paralelgia or Hemiplegia, Cancer, Moderate or severe liver disease, Metastatic solid tumor, HIV, Renal disease) were based on one inpatient, or two outpatient diagnoses (ICD-9 or ICD-10 codes) in the two years before index date. We defined outpatient infection diagnosis with any one outpatient code (ICD-9 or -10) in the 5 years prior to index date.

We identified outpatient antimicrobial prescription based on antimicrobial generic names from US National Drug codes.

England cohort

We defined smoking status based on a previously defined algorithm using primary care morbidity coding.^4^ We defined harmful alcohol use based on primary care recording before index date of: morbidity codes suggesting harmful or heavy alcohol use or a prescription for medications used to maintain abstinence. We defined primary care recorded infection with at least one relevant SNOMED-CT code recorded in primary care. We defined ethnicity based on a previously validated algorithm using primary care records.^5^

Care home residency was defined with primary care codes suggesting living in residential or nursing care home. We defined antibiotic prescriptions using primary care dictionary of medicines and device product codes representing all medications in Section 5.1 of the British National Formulary (excluding antituberculous and antilepromatous agents, and methenamine).

**Supplementary Appendix 3: Supplementary Tables**

| **Table S1 Description of sensitivity analyses conducted** | | | |
| --- | --- | --- | --- |
| **Sensitivity analysis** | **Cohort** | **Rationale** | **Methods** |
| Misclassification of severe-infection exposure | US and England | To explore the impact of potential misclassification of severe-infection exposure status | We repeated the fully adjusted models restricting to exposed individuals with evidence of outpatient or primary care infection diagnosis or pharmacy or primary care dispensed antimicrobial in the seven days before cohort entry, and their matched comparators |
| Missing VACS Index at index date | US | To explore the impact of missing VACS Index at index date | We included adults with missing but imputed VACS Index at index date in the fully adjusted linear regression models |
| Missing ethnicity data | England | To explore the impact of not adjusting for ethnicity (due to the proportion of missing data) | We additionally adjusted the fully adjusted linear regression models for ethnicity |
| Periods with potentially more complete data on eFI and other covariates | England | To explore the impact of more frequent recordings of eFI deficits and other covariates due to changes to the Quality Outcomes Framework (QOF) ^*^ indicators and other policy changes over time | We restricted the fully adjusted linear regression models to index dates from 2012 onwards and 2019 onwards. |
| ^*^ The Quality and Outcomes Framework (QOF) is a voluntary incentive program for GP practices in England, designed to reward high-quality care through a structured set of indicators and measures· From 2012 onwards the QOF indicators for osteoporosis, peripheral vascular disease, and the Prime Minister’s challenge were introduced. From 2014 onwards the Dementia Identification Scheme rewarded general practitioner practices for improving dementia detection rates. | | | |

| **Table S2. Supplementary characteristics of adults at index date** | | | | |
| --- | --- | --- | --- | --- |
|  | **US cohort** | | **England cohort** | |
|  | **No Severe infection,**  **n = 836,707** | **Severe infection,**  **n = 229,311** | **No Severe infection,**  **n = 1,208,374** | **Severe infection,**  **n = 243,743** |
| **Outpatient or primary care recorded infection**  **(7 days lookback)** | 11,884 (1·4%) | 170,185 (74%) | 7,866 (0·7%) | 58,019 (24%) |
| **Outpatient or primary care antibiotic prescription**  **(7 days lookback)** | 8,564 (1·0%) | 101,182 (44%) | 22,520 (1·9%) | 40,366 (17%) |
| **Myocardial infarction** | 28,347 (3·4%) | 22,452 (9·8%) | ·· | ·· |
| **Congestive heart failure** | 74,724 (8·9%) | 57,580 (25%) | ·· | ·· |
| **Peripheral vascular disease** | 101,919 (12%) | 59,772 (26%) | ·· | ·· |
| **Cerebrovascular disease** | 88,430 (11%) | 48,128 (21%) | 109,111 (9·0%) | 34,706 (14%) |
| **Chronic Obstructive Pulmonary Disease** | 176,662 (21%) | 90,308 (39%) | ·· | ·· |
| **Rheumatologic disease** | 17,324 (2·1%) | 7,504 (3·3%) | 41,633 (3·4%) | 11,919 (4·9%) |
| **Peptic ulcer disease** | 11,368 (1·4%) | 7,119 (3·1%) | ·· | ·· |
| **Mild liver disease** | 31,956 (3·8%) | 17,173 (7·5%) | ·· | ·· |
| **Paralelgia or Hemiplegia** | 118,539 (14%) | 61,392 (27%) | ·· | ·· |
| **Cancer** | 5,537 (0·7%) | 6,115 (2·7%) | ·· | ·· |
| **Moderate or severe liver disease** | 135,016 (16%) | 63,957 (28%) | ·· | ·· |
| **Metastatic solid tumor** | 3,705 (0·4%) | 3,463 (1·5%) | ·· | ·· |
| **HIV** | 9,012 (1·1%) | 12,269 (5·4%) | ·· | ·· |
| **Renal disease** | 1,130 (0·1%) | 590 (0·3%) | ·· | ·· |
| Data are n (%).  All percentages were calculated with the total number in the respective column headers as denominators.  Percentages may not add up to 100 due to rounding.  IQR = Interquartile Range, SD = Standard Deviation. | | | | |

| **Table S3. US cohort: Linear regression for conditional mean differences in VACS Index at follow-up years 1 to 5 between adults with and without severe infection at index date** | | | | | | | | | | |
| --- | --- | --- | --- | --- | --- | --- | --- | --- | --- | --- |
|  | **Year 1** | | **Year 2** | | **Year 3** | | **Year 4** | | **Year 5** | |
|  | **N** | **Conditional mean difference in VACS Index (95% CI)** | **N** | **Conditional mean difference in VACS Index (95% CI)** | **N** | **Conditional mean difference in VACS Index (95% CI)** | **N** | **Conditional mean difference in VACS Index (95% CI)** | **N** | **Conditional mean difference in VACS Index (95% CI)** |
| Adjusted for age, sex, race and ethnicity, and VACS Index at index date | 900,882 | 2·8 (2·7, 2·8) | 700,860 | 1·1 (1·1, 1·2) | 556,952 | 1·1 (1·0, 1·1) | 444,073 | 1·1 (1·0, 1·1) | 346,620 | 1·1 (1·1, 1·2) |
| Further adjusted for index of area deprivation and area of residence at index date | 879,600 | 2·7 (2·6, 2·7) | 692,097 | 1·1 (1·0, 1·1) | 553,411 | 1·0 (1·0, 1·1) | 442,835 | 1·0 (1·0, 1·1) | 346,011 | 1·1 (1·1, 1·2) |
| Further adjusted for smoking status and alcohol use and use disorder at index date | 856,672 | 2·5 (2·5, 2·6) | 675,222 | 1·0 (0·9, 1·0) | 540,426 | 0·9 (0·9, 1·0) | 432,644 | 0·9 (0·8, 1·0) | 338,052 | 1·0 (0·9, 1·1) |
| Further adjusted for CCI at index date and outpatient recorded infection 5 years before index date (fully adjusted model) | 856,672 | 2·0 (1·9, 2·0) | 675,222 | 0·4 (0·3, 0·4) | 540,426 | 0·3 (0·2, 0·4) | 432,644 | 0·3 (0·2, 0·3) | 338,052 | 0·4 (0·3, 0·5) |
| **Sensitivity analysis 1:** Added adults with missing but imputed VACS Index at index date· | 915384 | 2·0 (1·9, 2·0) | 730542 | 0·4 (0·3, 0·4) | 590439 | 0·3 (0·2, 0·4) | 477632 | 0·3 (0·2, 0·4) | 377153 | 0·4 (0·3, 0·5) |
| **Sensitivity analysis 2:** Restricted analysis to matched sets where the exposed adult had records indicating community acquired infection exposure 7 days before index date· |  |  |  |  |  |  |  |  |  |  |
| a. Outpatient infection diagnosis | 652,436 | 2·0 (1·9, 2·1) | 508,083 | 0·5 (0·4, 0·5) | 399,684 | 0·3 (0·3, 0·4) | 315,119 | 0·4 (0·3, 0·4) | 241,838 | 0·5 (0·3, 0·6) |
| b. Outpatient dispensed antimicrobial | 390,715 | 1·8 (1·7, 1·9) | 319,561 | 0·7 (0·6, 0·7) | 265,471 | 0·6 (0·5, 0·7) | 216,326 | 0·5 (0·4, 0·6) | 171,067 | 0·7 (0·6, 0·8) |
| c. Having both a· and b· | 290,775 | 1·8 (1·7, 1·9) | 236,696 | 0·8 (0·7, 0·8) | 195,048 | 0·6 (0·5, 0·8) | 157,041 | 0·6 (0·4, 0·7) | 122,042 | 0·7 (0·6, 0·9) |
| CCI = Charlson Comorbidity Index, CI = Confidence Interval, VACS = Veterans Aging Cohort Study. | | | | | | | | | | |

| **Table S4. England cohort: Linear regression for conditional mean differences in Electronic Frailty Index at follow-up years 1 to 5 between adults with and without severe infection at index date** | | | | | | | | | | |
| --- | --- | --- | --- | --- | --- | --- | --- | --- | --- | --- |
|  | Year 1 | | Year 2 | | Year 3 | | Year 4 | | Year 5 | |
|  | N | Conditional mean difference in eFI (95% CI) | N | Conditional mean difference in eFI (95% CI) | N | Conditional mean difference in eFI (95% CI) | N | Conditional mean difference in eFI (95% CI) | N | Conditional mean difference in eFI (95% CI) |
| Adjusted for age, sex, and eFI at index date | 1,452,117 | 0·006 (0·006, 0·006) | 1,191,961 | 0·011 (0·011, 0·012) | 995,227 | 0·014 (0·013, 0·014) | 824,075 | 0·015 (0·015, 0·016) | 678,628 | 0·017 (0·016, 0·017) |
| Further adjusted for index of multiple deprivation quintiles at index date | 1,452,117 | 0·006 (0·006, 0·006) | 1,191,961 | 0·011 (0·011, 0·012) | 995,227 | 0·014 (0·013, 0·014) | 824,075 | 0·015 (0·015, 0·016) | 678,628 | 0·017 (0·016, 0·017) |
| Further adjusted for smoking and harmful alcohol use at index date | 1,430,375 | 0·006 (0·006, 0·006) | 1,174,447 | 0·011 (0·011, 0·011) | 980,792 | 0·013 (0·013, 0·014) | 812,191 | 0·015 (0·015, 0·015) | 668,816 | 0·016 (0·016, 0·017) |
| Further adjusted for primary care recorded infection 5 years before index date (fully adjusted model) | 1,430,375 | 0·005 (0·005, 0·006) | 1,174,447 | 0·011 (0·010, 0·011) | 980,792 | 0·012 (0·012, 0·013) | 812,191 | 0·014 (0·013, 0·014) | 668,816 | 0·015 (0·014, 0·015) |
| Sensitivity analysis 1: Further adjusted for ethnicity | 848,494 | 0·006 (0·006, 0·006) | 682,204 | 0·011 (0·011, 0·011) | 553,972 | 0·013 (0·013, 0·013) | 443,350 | 0·015 (0·014, 0·015) | 351,303 | 0·016 (0·015, 0·017) |
| Sensitivity analysis 2: Restricted analysis to matched sets where the exposed adult had records indicating community acquired infection exposure 7 days before index date· |  |  |  |  |  |  |  |  |  |  |
| Primary care infection diagnosis | 341,255 | 0·006 (0·006, 0·007) | 279,489 | 0·011 (0·011, 0·012) | 230,636 | 0·013 (0·013, 0·014) | 189,128 | 0·015 (0·014, 0·016) | 153,912 | 0·016 (0·015, 0·017) |
| Primary care dispensed antimicrobial | 237,320 | 0·006 (0·005, 0·006) | 195,443 | 0·011 (0·011, 0·012) | 164,304 | 0·013 (0·013, 0·014) | 136,822 | 0·015 (0·014, 0·016) | 112,652 | 0·017 (0·015, 0·018) |
| Having both a· and b· | 237,320 | 0·006 (0·005, 0·006) | 195,443 | 0·011 (0·011, 0·012) | 164,304 | 0·013 (0·013, 0·014) | 136,822 | 0·015 (0·014, 0·016) | 112,652 | 0·017 (0·015, 0·018) |
| Sensitivity analysis 3: Restricted analysis to Quality Outcomes Framework periods· |  |  |  |  |  |  |  |  |  |  |
| 2012 to 2019 | 786,931 | 0·006 (0·006, 0·007) | 613,504 | 0·012 (0·012, 0·012) | 476,387 | 0·014 (0·014, 0·015) | 357,816 | 0·016 (0·016, 0·017) | 259,310 | 0·018 (0·017, 0·018) |
| 2014 to 2019 | 586,399 | 0·007 (0·006, 0·007) | 438,312 | 0·012 (0·012, 0·012) | 319,125 | 0·014 (0·014, 0·015) | 215,620 | 0·016 (0·016, 0·017) | 130,613 | 0·018 (0·017, 0·019) |
| CI = confidence interval, eFI = Electronic Frailty Index. | | | | | | | | | | |

| **Table S5. US cohort: Cohort characteristics at index date including adults with missing VACS Index** | | | | |
| --- | --- | --- | --- | --- |
|  | **Cohort including adults with missing VACS Index at index date** | | **Cohort of adults with available VACS Index at index date**  **(main analysis, Table1)** | |
|  | **No Severe infection**,  n = 987,894 | **Severe infection**,  n = 243,758 | **No Severe infection,**  n = 836,707 | **Severe infection,**  n = 229,311 |
| **Age, years** |  |  |  |  |
| Median (IQR) | 74 (69, 81) | 74 (70, 82) | 74 (69, 81) | 74 (70, 82) |
| 65-70 | 282,262 (29%) | 65,371 (27%) | 236,333 (28%) | 61,236 (27%) |
| 71-80 | 437,379 (44%) | 105,866 (43%) | 376,158 (45%) | 100,188 (44%) |
| 81-90 | 224,492 (23%) | 57,956 (24%) | 188,061 (22%) | 54,381 (24%) |
| >90 | 43,761 (4·4%) | 14,565 (6·0%) | 36,155 (4·3%) | 13,506 (5·9%) |
| **Sex** |  |  |  |  |
| Female | 18,803 (1·9%) | 5,093 (2·1%) | 15,984 (1·9%) | 4,799 (2·1%) |
| Male | 969,091 (98%) | 238,665 (98%) | 820,723 (98%) | 224,512 (98%) |
| **Race and ethnicity** |  |  |  |  |
| White | 739,660 (75%) | 179,751 (74%) | 625,769 (75%) | 169,253 (74%) |
| Black | 135,633 (14%) | 33,634 (14%) | 115,866 (14%) | 31,597 (14%) |
| Hispanic | 66,631 (6·7%) | 17,156 (7·0%) | 58,092 (6·9%) | 16,340 (7·1%) |
| Other | 15,845 (1·6%) | 4,088 (1·7%) | 13,548 (1·6%) | 3,839 (1·7%) |
| Asian | 2,498 (0·3%) | 747 (0·3%) | 1,945 (0·2%) | 688 (0·3%) |
| Missing | 27,627 (2·8%) | 8,382 (3·4%) | 21,487 (2·6%) | 7,594 (3·3%) |
| **Area of residence** |  |  |  |  |
| Rural | 370,773 (38%) | 81,247 (33%) | 317,086 (38%) | 76,678 (33%) |
| Urban | 617,121 (62%) | 162,511 (67%) | 519,621 (62%) | 152,633 (67%) |
| **Area Deprivation Index (quintiles)** |  |  |  |  |
| First, least deprived | 176,236 (18%) | 41,536 (17%) | 142,475 (17%) | 38,306 (17%) |
| Second | 190,663 (19%) | 43,701 (18%) | 158,485 (19%) | 40,817 (18%) |
| Third | 201,363 (20%) | 47,015 (19%) | 170,598 (20%) | 44,299 (19%) |
| Fourth | 202,911 (21%) | 50,115 (21%) | 175,221 (21%) | 47,502 (21%) |
| Fifth | 200,249 (20%) | 51,426 (21%) | 175,719 (21%) | 48,944 (21%) |
| Missing | 16,472 (1·7%) | 9,965 (4·1%) | 14,209 (1·7%) | 9,443 (4·1%) |
| **Alcohol use and use disorder** |  |  |  |  |
| Abstinent without AUD | 504,596 (51%) | 159,311 (65%) | 450,561 (54%) | 151,018 (66%) |
| Abstinent with AUD | 7,774 (0·8%) | 5,500 (2·3%) | 7,352 (0·9%) | 5,387 (2·3%) |
| Lower-risk consumption | 293,285 (30%) | 49,922 (20%) | 255,924 (31%) | 46,568 (20%) |
| Moderate-risk consumption | 88,826 (9·0%) | 12,334 (5·1%) | 76,440 (9·1%) | 11,367 (5·0%) |
| High-risk consumption | 28,195 (2·9%) | 10,128 (4·2%) | 25,898 (3·1%) | 9,646 (4·2%) |
| Missing | 65,218 (6·6%) | 6,563 (2·7%) | 20,532 (2·5%) | 5,325 (2·3%) |
| **Smoking status** |  |  |  |  |
| Current | 207,090 (21%) | 67,624 (28%) | 179,299 (21%) | 63,761 (28%) |
| Former | 464,803 (47%) | 106,364 (44%) | 389,947 (47%) | 99,994 (44%) |
| Never | 309,294 (31%) | 68,086 (28%) | 262,810 (31%) | 64,111 (28%) |
| Missing | 6,707 (0·7%) | 1,684 (0·7%) | 4,651 (0·6%) | 1,445 (0·6%) |
| **Charlson Comorbidity Index** |  |  |  |  |
| 0 | 543,279 (55%) | 59,369 (24%) | 429,270 (51%) | 51,899 (23%) |
| 1 | 187,612 (19%) | 41,275 (17%) | 167,536 (20%) | 38,595 (17%) |
| 2 | 110,228 (11%) | 37,352 (15%) | 101,011 (12%) | 35,591 (16%) |
| 3+ | 146,775 (15%) | 105,762 (43%) | 138,890 (17%) | 103,226 (45%) |
| **Myocardial infarction** | 30,244 (3·1%) | 22,950 (9·4%) | 28,347 (3·4%) | 22,452 (9·8%) |
| **Congestive heart failure** | 79,903 (8·1%) | 59,202 (24%) | 74,724 (8·9%) | 57,580 (25%) |
| **Peripheral vascular disease** | 110,099 (11%) | 61,710 (25%) | 101,919 (12%) | 59,772 (26%) |
| **Cerebrovascular disease** | 95,933 (9·7%) | 49,775 (20%) | 88,430 (11%) | 48,128 (21%) |
| **Dementia** | 29,884 (3·0%) | 22,754 (9·3%) | 27,676 (3·3%) | 21,923 (9·6%) |
| **Chronic Obstructive Pulmonary Disease** | 192,888 (20%) | 93,649 (38%) | 176,662 (21%) | 90,308 (39%) |
| **Rheumatologic disease** | 18,559 (1·9%) | 7,693 (3·2%) | 17,324 (2·1%) | 7,504 (3·3%) |
| **Peptic ulcer disease** | 12,237 (1·2%) | 7,257 (3·0%) | 11,368 (1·4%) | 7,119 (3·1%) |
| **Mild liver disease** | 33,362 (3·4%) | 17,504 (7·2%) | 31,956 (3·8%) | 17,173 (7·5%) |
| **Diabetes mellitus** | 345,738 (35%) | 117,176 (48%) | 311,124 (37%) | 111,911 (49%) |
| **Paralelgia or Hemiplegia** | 128,486 (13%) | 63,656 (26%) | 118,539 (14%) | 61,392 (27%) |
| **Cancer** | 5,901 (0·6%) | 6,325 (2·6%) | 5,537 (0·7%) | 6,115 (2·7%) |
| **Moderate or severe liver disease** | 147,469 (15%) | 65,843 (27%) | 135,016 (16%) | 63,957 (28%) |
| **Metastatic solid tumor** | 3,827 (0·4%) | 3,507 (1·4%) | 3,705 (0·4%) | 3,463 (1·5%) |
| **HIV** | 9,263 (0·9%) | 12,417 (5·1%) | 9,012 (1·1%) | 12,269 (5·4%) |
| **Renal disease** | 1,368 (0·1%) | 683 (0·3%) | 1,130 (0·1%) | 590 (0·3%) |
| **Outpatient diagnosed infection (5-year lookback)** | 363,970 (37%) | 141,132 (58%) | 28,347 (3·4%) | 22,452 (9·8%) |
| **VACS Index** |  |  |  |  |
| Mean (SD) | 48 (11) | 57 (14) | 48 (11) | 57 (14) |
| Missing | 151,187 | 14,447 | ·· | ·· |
| Data are n (%), mean (SD), or median (IQR). All percentages were calculated with the total number in the respective column headers as denominators. Percentages may not add up to 100 due to rounding.  AUD = Alcohol Use Disorder, IQR = Interquartile Range, SD = Standard Deviation, VACS = Veterans Aging Cohort Study. | | | | |

| **Table S6. US cohort: VACS Index during follow-up in cohort including adults with missing VACS Index at index date** | | | | | | | | | | | | |
| --- | --- | --- | --- | --- | --- | --- | --- | --- | --- | --- | --- | --- |
|  | **Overall** | | **Year 1** | | **Year 2** | | **Year 3** | | **Year 4** | | **Year 5** | |
|  | Comparator | Severe infection | Comparator | Severe infection | Comparator | Severe infection | Comparator | Severe infection | Comparator | Severe infection | Comparator | Severe infection |
| Active number of adults | 987,894 | 243,758 | 987,834 | 243,758 | 887,352 | 185,384 | 743,728 | 144,693 | 610,133 | 112,536 | 489,736 | 86,858 |
| Median (IQR) follow-up, years | 4·0 (2·0, 6·5) | 2·7 (1·1, 5·2) |  |  |  |  |  |  |  |  |  |  |
| Died during year |  |  | <50 (<0·1%) | 0 (0%) | 16,599 (1·7%) | 30,073 (12%) | 38,068 (3·9%) | 45,453 (19%) | 57,727 (5·8%) | 56,703 (23%) | 74,899 (7·6%) | 65,313 (27%) |
| Censored for other reasons during year |  |  | >50 (<0·1%) | 0 (0%) | 83,943 (8·5%) | 28,301 (12%) | 206,098 (21%) | 53,612 (22%) | 320,034 (32%) | 74,519 (31%) | 423,259 (43%) | 91,587 (38%) |
| Mean (SD) VACS Index during year |  |  | 50 (12) | 60 (16) | 50 (11) | 57 (14) | 51 (11) | 57 (14) | 52 (11) | 57 (13) | 52 (11) | 57 (13) |
| Data are n (%), mean (SD), or median (IQR).  All percentages were calculated with the population at index date as the denominator.  Censoring for other reasons included study end or last VA visit.  To prevent disclosure, we have used primary suppression to directly suppress cells with small counts (i.e., <50) and secondary suppression to suppress additional cells that do not have small counts themselves, but which need to be suppressed to protect the values in the primarily suppressed cells.  IQR = Interquartile Range, SD = Standard Deviation, VA = Department of Veterans’ Affairs, VACS = Veterans Aging Cohort Study. | | | | | | | | | | | | |

| **Table S7. US cohort: Stratified linear regression for conditional mean differences in VACS Index at follow-up years 1 to 5 between adults with and without severe infection at index date** | | | | | | | | | | |
| --- | --- | --- | --- | --- | --- | --- | --- | --- | --- | --- |
|  | **Year 1** | | **Year 2** | | **Year 3** | | **Year 4** | | **Year 5** | |
|  | **N** | **Conditional mean difference in VACS Index (95% CI)** | **N** | **Conditional mean difference in VACS Index (95% CI)** | **N** | **Conditional mean difference in VACS Index (95% CI)** | **N** | **Conditional mean difference in VACS Index (95% CI)** | **N** | **Conditional mean difference in VACS Index (95% CI)** |
| **Age** |  |  |  |  |  |  |  |  |  |  |
| 65-70 | 239,559 | 1·3 (1·2, 1·4) | 206,823 | 0·0 (-0·1, 0·1) | 180,163 | -0·1 (-0·2, 0·0) | 156,454 | -0·1 (-0·2, 0·0) | 132,462 | 0·0 (-0·1, 0·2) |
| 71-80 | 390,789 | 1·8 (1·7, 1·9) | 303,649 | 0·3 (0·2, 0·4) | 236,552 | 0·3 (0·2, 0·4) | 184,121 | 0·3 (0·2, 0·4) | 138,908 | 0·5 (0·3, 0·6) |
| 81-90 | 188,003 | 2·7 (2·6, 2·8) | 141,026 | 0·9 (0·7, 1·0) | 108,291 | 0·7 (0·6, 0·9) | 82,247 | 0·7 (0·5, 0·9) | 60,671 | 0·8 (0·6, 1·0) |
| >90 | 38,321 | 3·8 (3·5, 4·0) | 23,724 | 1·7 (1·4, 2·0) | 15,420 | 1·5 (1·1, 1·9) | 9,822 | 1·4 (0·9, 2·0) | 6,011 | 2·0 (1·2, 2·7) |
| **Index of area deprivation quintile** |  |  |  |  |  |  |  |  |  |  |
| 1, least deprived | 143,158 | 2·0 (1·9, 2·1) | 112,843 | 0·4 (0·2, 0·5) | 90,482 | 0·3 (0·2, 0·5) | 72,902 | 0·4 (0·2, 0·6) | 57,404 | 0·6 (0·4, 0·8) |
| 2 | 161,098 | 2·1 (2·0, 2·2) | 126,831 | 0·4 (0·3, 0·5) | 101,813 | 0·3 (0·1, 0·5) | 81,390 | 0·3 (0·2, 0·5) | 63,170 | 0·3 (0·1, 0·6) |
| 3 | 176,397 | 2·0 (1·9, 2·1) | 139,463 | 0·4 (0·3, 0·6) | 111,801 | 0·4 (0·3, 0·6) | 89,327 | 0·3 (0·2, 0·5) | 69,315 | 0·4 (0·2, 0·6) |
| 4 | 186,452 | 1·8 (1·7, 1·9) | 146,828 | 0·4 (0·3, 0·6) | 117,365 | 0·2 (0·1, 0·4) | 93,598 | 0·2 (0·1, 0·4) | 73,085 | 0·4 (0·2, 0·6) |
| 5, most deprived | 189,567 | 1·9 (1·8, 2·0) | 149,257 | 0·2 (0·1, 0·3) | 118,965 | 0·2 (0, 0·3·0) | 95,427 | 0·0 (-0·1, 0·2) | 75,078 | 0·2 (0·0, 0·4) |
| **Dementia status** |  |  |  |  |  |  |  |  |  |  |
| No | 816,831 | 1·8 (1·8, 1·9) | 652,206 | 0·3 (0·2, 0·3) | 525,944 | 0·2 (0·2, 0·3) | 423,489 | 0·2 (0·1, 0·3) | 332,533 | 0·4 (0·3, 0·4) |
| Yes | 39,841 | 3·1 (2·9, 3·3) | 23,016 | 1·5 (1·2, 1·8) | 14,482 | 1·0 (0·6, 1·4) | 9,155 | 1·3 (0·8, 1·8) | 5,519 | 1·0 (0·4, 1·6) |
| **Diabetes status** |  |  |  |  |  |  |  |  |  |  |
| No | 508,496 | 2·2 (2·1, 2·3) | 405,179 | 0·4 (0·3, 0·5) | 327,095 | 0·3 (0·2, 0·4) | 264,518 | 0·2 (0·1, 0·3) | 209,017 | 0·3 (0·1, 0·4) |
| Yes | 348,176 | 1·7 (1·6, 1·7) | 270,043 | 0·3 (0·2, 0·4) | 213,331 | 0·3 (0·2, 0·4) | 168,126 | 0·4 (0·2, 0·5) | 129,035 | 0·5 (0·4, 0·6) |
| **Charlson Comorbidity Index** |  |  |  |  |  |  |  |  |  |  |
| 0 | 374,684 | 2·0 (1·9, 2·1) | 313,553 | 0·5 (0·4, 0·6) | 259,906 | 0·4 (0·3, 0·5) | 215,693 | 0·3 (0·2, 0·4) | 174,038 | 0·4 (0·3, 0·6) |
| 1 | 165,668 | 2·0 (1·9, 2·1) | 133,450 | 0·5 (0·4, 0·6) | 108,012 | 0·4 (0·3, 0·6) | 86,929 | 0·3 (0·1, 0·4) | 67,917 | 0·3 (0·1, 0·5) |
| 2 | 111,859 | 1·9 (1·8, 2·1) | 87,098 | 0·4 (0·3, 0·6) | 69,197 | 0·3 (0·1, 0·4) | 54,247 | 0·4 (0·2, 0·6) | 41,691 | 0·4 (0·1, 0·6) |
| 3+ | 204,461 | 2·2 (2·1, 2·3) | 141,121 | 0·5 (0·4, 0·6) | 103,311 | 0·4 (0·3, 0·6) | 75,775 | 0·5 (0·3, 0·6) | 54,406 | 0·7 (0·5, 0·9) |
| **Pathogen of severe infection exposure** |  |  |  |  |  |  |  |  |  |  |
| Bacterial | 676,159 | 2·0 (1·9, 2·0) | 543,820 | 0·3 (0·2, 0·3) | 448,989 | 0·1 (0·1, 0·2) | 363,853 | 0·1 (0·0, 0·2) | 285,855 | 0·2 (0·1, 0·3) |
| Viral | 126,370 | 1·6 (1·5, 1·7) | 88,593 | 0·5 (0·4, 0·7) | 56,677 | 0·9 (0·7, 1·0) | 42,040 | 1·0 (0·7, 1·2) | 32,750 | 1·0 (0·8, 1·3) |
| Fungal | 5,182 | 2·2 (1·4, 3·0) | 4,270 | -0·4 (-1·3, 0·5) | 3,564 | -0·2 (-1·2, 0·8) | 3,014 | -1·2 (-2·4, 0·1) | 2,375 | 0·6 (-1, 2·1·0) |
| Parasitic | 1,085 | 0·8 (-0·5, 2·0) | 918 | -0·3 (-1·8, 1·1) | 783 | 0·5 (-1·4, 2·4) | 681 | -0·6 (-2·5, 1·3) | 592 | -0·3 (-2·3, 1·8) |
| **Type of severe infection exposure** |  |  |  |  |  |  |  |  |  |  |
| Sepsis | 172,882 | 3·0 (2·9, 3·1) | 130,539 | 0·6 (0·5, 0·8) | 101,749 | 0·5 (0·3, 0·7) | 77,322 | 0·5 (0·2, 0·7) | 55,649 | 0·4 (0·1, 0·6) |
| Urinary Tract | 123,013 | 2·6 (2·4, 2·7) | 98,322 | 0·8 (0·7, 1·0) | 80,637 | 0·6 (0·4, 0·8) | 65,159 | 0·5 (0·2, 0·7) | 52,023 | 0·5 (0·3, 0·8) |
| Skin and soft tissue | 135,099 | 1·7 (1·6, 1·8) | 112,981 | 0·8 (0·7, 0·9) | 96,224 | 0·8 (0·6, 0·9) | 80,430 | 1·1 (0·9, 1·2) | 65,627 | 1·2 (1·0, 1·4) |
| Meningoencephalitis | 1,794 | 0·4 (-0·8, 1·5) | 1,566 | -0·5 (-1·6, 0·6) | 1,382 | -0·4 (-1·7, 1·0) | 1,171 | -0·2 (-1·6, 1·2) | 978 | 0·3 (-1·2, 1·9) |
| Lower respiratory tract | 150,299 | 2·9 (2·7, 3·0) | 120,784 | 1·5 (1·3, 1·6) | 99,735 | 1·5 (1·3, 1·7) | 78,974 | 1·3 (1·1, 1·5) | 59,397 | 1·5 (1·3, 1·8) |
| Gastroenteritis | 676,159 | 2·0 (1·9, 2·0) | 543,820 | 0·3 (0·2, 0·3) | 448,989 | 0·1 (0·1, 0·2) | 363,853 | 0·1 (0·0, 0·2) | 285,855 | 0·2 (0·1, 0·3) |
| All models are stratified from the fully adjusted model in Table S3.  Interaction p-values: Age: Years 1-5: < 0·0001; Sex: Year 1: < 0·0001; Year 2: 0·0001; Year 3: 0·008; Year 4: 0·022; Year 5: 0·18; Index of area deprivation quintile: Year 1: 0·102; Year 2: 0·006; Years 3-5: < 0·0001; Dementia status: Years 1-5: < 0·0001; Diabetes status: Year 1: 0·495, Year 2: 0·0001, Years 3-5: < 0·0001; Charlson Comorbidity Index: Years 1-3: < 0·0001, Year 4: 0·024, Year 5: 0·68; Pathogen of severe infection exposure: Years 1-5: > 0·05, Type of severe infection exposure: Years 1-5: < 0·0001.  CI = Confidence Interval, VACS = Veterans Aging Cohort Study. | | | | | | | | | | |

| **Table S8. England cohort: Stratified linear regression for conditional mean differences in Electronic Frailty Index at follow-up years 1 to 5 between adults with and without severe infection at index date** | | | | | | | | | | |
| --- | --- | --- | --- | --- | --- | --- | --- | --- | --- | --- |
|  | **Year 1** | | **Year 2** | | **Year 3** | | **Year 4** | | **Year 5** | |
|  | **N** | **Conditional mean difference in eFI**  **(95% CI)** | **N** | **Conditional mean difference in eFI (95% CI)** | **N** | **Conditional mean difference in eFI (95% CI)** | **N** | **Conditional mean difference in eFI (95% CI)** | **N** | **Conditional mean difference in eFI (95% CI)** |
| **Age** |  |  |  |  |  |  |  |  |  |  |
| 65-70 | 400,035 | 0·006 (0·005, 0·006) | 346,955 | 0·009 (0·009, 0·009) | 302,469 | 0·011 (0·011, 0·011) | 260,900 | 0·012 (0·012, 0·013) | 223,064 | 0·013 (0·013, 0·014) |
| 71-80 | 526,874 | 0·006 (0·006, 0·007) | 447,832 | 0·011 (0·011, 0·012) | 384,592 | 0·014 (0·013, 0·014) | 327,633 | 0·015 (0·015, 0·016) | 277,700 | 0·016 (0·016, 0·017) |
| 81-90 | 413,206 | 0·005 (0·004, 0·005) | 321,728 | 0·011 (0·011, 0·012) | 255,225 | 0·012 (0·012, 0·013) | 198,937 | 0·013 (0·013, 0·014) | 152,363 | 0·014 (0·013, 0·015) |
| >90 | 90,260 | 0·001 (0·001, 0·002) | 57,932 | 0·008 (0·007, 0·009) | 38,506 | 0·009 (0·007, 0·010) | 24,721 | 0·008 (0·006, 0·011) | 15,689 | 0·010 (0·007, 0·013) |
| **Sex** |  |  |  |  |  |  |  |  |  |  |
| Female | 786,195 | 0·005 (0·005, 0·005) | 644,210 | 0·010 (0·010, 0·011) | 537,122 | 0·012 (0·012, 0·013) | 444,413 | 0·014 (0·013, 0·014) | 365,616 | 0·015 (0·014, 0·015) |
| Male | 644,180 | 0·006 (0·006, 0·006) | 530,237 | 0·011 (0·011, 0·011) | 443,670 | 0·013 (0·012, 0·013) | 367,778 | 0·014 (0·013, 0·014) | 303,200 | 0·015 (0·014, 0·016) |
| **Index of multiple deprivation quintiles** |  |  |  |  |  |  |  |  |  |  |
| 1, least deprived | 319,335 | 0·005 (0·005, 0·005) | 263,806 | 0·010 (0·009, 0·010) | 221,379 | 0·011 (0·011, 0·012) | 183,738 | 0·012 (0·011, 0·013) | 151,965 | 0·013 (0·012, 0·014) |
| 2 | 310,005 | 0·005 (0·005, 0·005) | 255,852 | 0·010 (0·010, 0·010) | 213,986 | 0·011 (0·011, 0·012) | 177,213 | 0·013 (0·012, 0·013) | 146,193 | 0·014 (0·013, 0·015) |
| 3 | 289,593 | 0·006 (0·005, 0·006) | 237,340 | 0·011 (0·010, 0·011) | 198,069 | 0·012 (0·012, 0·013) | 163,675 | 0·014 (0·013, 0·014) | 134,562 | 0·015 (0·014, 0·016) |
| 4 | 269,290 | 0·006 (0·005, 0·006) | 220,166 | 0·011 (0·011, 0·012) | 183,257 | 0·014 (0·013, 0·014) | 151,813 | 0·015 (0·015, 0·016) | 124,850 | 0·016 (0·015, 0·017) |
| 5, most deprived | 242,152 | 0·006 (0·005, 0·006) | 197,283 | 0·012 (0·011, 0·012) | 164,101 | 0·014 (0·013, 0·014) | 135,752 | 0·015 (0·015, 0·016) | 111,246 | 0·016 (0·015, 0·017) |
| **Care home residency** |  |  |  |  |  |  |  |  |  |  |
| No | 1,399,736 | 0·005 (0·005, 0·006) | 1,157,457 | 0·011 (0·010, 0·011) | 971,301 | 0·012 (0·012, 0·013) | 806,953 | 0·014 (0·013, 0·014) | 665,896 | 0·015 (0·014, 0·015) |
| Yes | 30,639 | 0·001 (0·000, 0·002) | 16,990 | 0·007 (0·006, 0·009) | 9,491 | 0·010 (0·007, 0·012) | 5,238 | 0·011 (0·007, 0·015) | 2,920 | 0·014 (0·009, 0·020) |
| **Dementia status** |  |  |  |  |  |  |  |  |  |  |
| No | 1,340,811 | 0·006 (0·006, 0·006) | 1,120,127 | 0·011 (0·010, 0·011) | 946,664 | 0·012 (0·012, 0·013) | 791,080 | 0·014 (0·013, 0·014) | 656,047 | 0·015 (0·014, 0·015) |
| Yes | 89,564 | 0·002 (0·001, 0·002) | 54,320 | 0·008 (0·008, 0·009) | 34,128 | 0·009 (0·008, 0·011) | 21,111 | 0·012 (0·010, 0·013) | 12,769 | 0·011 (0·008, 0·014) |
| **Diabetes status** |  |  |  |  |  |  |  |  |  |  |
| No | 1,156,699 | 0·005 (0·005, 0·005) | 959,274 | 0·010 (0·010, 0·010) | 807,713 | 0·012 (0·011, 0·012) | 674,215 | 0·013 (0·012, 0·013) | 559,557 | 0·014 (0·013, 0·014) |
| Yes | 273,676 | 0·006 (0·005, 0·006) | 215,173 | 0·012 (0·011, 0·012) | 173,079 | 0·014 (0·013, 0·014) | 137,976 | 0·016 (0·015, 0·016) | 109,259 | 0·017 (0·016, 0·018) |
| **Pathogen of severe infection exposure** |  |  |  |  |  |  |  |  |  |  |
| Bacterial | 1,330,379 | 0·005 (0·005, 0·006) | 1,091,221 | 0·011 (0·011, 0·011) | 910,887 | 0·013 (0·012, 0·013) | 753,775 | 0·014 (0·013, 0·014) | 620,499 | 0·015 (0·014, 0·015) |
| Viral | 82,078 | 0·006 (0·005, 0·006) | 68,765 | 0·009 (0·008, 0·010) | 57,869 | 0·011 (0·010, 0·012) | 48,351 | 0·013 (0·012, 0·015) | 40,111 | 0·014 (0·013, 0·016) |
| Fungal | 13,349 | 0·005 (0·003, 0·006) | 11,088 | 0·008 (0·006, 0·010) | 9,367 | 0·010 (0·007, 0·013) | 7,907 | 0·013 (0·010, 0·017) | 6,478 | 0·015 (0·011, 0·019) |
| Parasitic | 3,918 | 0·006 (0·004, 0·008) | 2,860 | 0·009 (0·005, 0·013) | 2,247 | 0·011 (0·006, 0·016) | 1,791 | 0·013 (0·007, 0·020) | 1,421 | 0·016 (0·007, 0·024) |
| **Type of severe infection exposure** |  |  |  |  |  |  |  |  |  |  |
| Sepsis | 160,822 | 0·003 (0·003, 0·004) | 119,390 | 0·014 (0·013, 0·014) | 89,320 | 0·015 (0·014, 0·016) | 62,161 | 0·016 (0·014, 0·017) | 48,214 | 0·017 (0·015, 0·019) |
| Urinary Tract | 466,994 | 0·007 (0·006, 0·007) | 378,724 | 0·013 (0·012, 0·013) | 315,212 | 0·015 (0·014, 0·015) | 262,249 | 0·016 (0·015, 0·016) | 213,124 | 0·017 (0·016, 0·018) |
| Skin and soft tissue | 217,758 | 0·005 (0·005, 0·005) | 183,287 | 0·008 (0·008, 0·009) | 155,974 | 0·010 (0·010, 0·011) | 131,675 | 0·012 (0·011, 0·013) | 110,367 | 0·013 (0·012, 0·014) |
| Meningoencephalitis | 5,188 | 0·006 (0·004, 0·008) | 4,389 | 0·012 (0·009, 0·015) | 3,794 | 0·015 (0·011, 0·019) | 3,276 | 0·020 (0·015, 0·025) | 2,748 | 0·020 (0·014, 0·027) |
| Lower respiratory tract | 225,249 | 0·007 (0·007, 0·008) | 186,415 | 0·014 (0·014, 0·015) | 156,704 | 0·018 (0·017, 0·018) | 131,340 | 0·020 (0·019, 0·021) | 107,765 | 0·022 (0·021, 0·024) |
| Gastroenteritis | 65,155 | 0·004 (0·003, 0·004) | 54,181 | 0·009 (0·009, 0·010) | 46,202 | 0·012 (0·011, 0·013) | 39,079 | 0·013 (0·012, 0·015) | 32,510 | 0·014 (0·012, 0·016) |
| All models are stratified from the fully adjusted model in Table S4.  Interaction p-values: Age: Years 1-5: < 0·0001; Sex: Year 1-2: < 0·0001; Year 3: 0·001; Year 4: 0·039; Year 5: 0·80; Index of multiple deprivation quintile: Year 1-5: < 0·0001; Care home residency: Year 1: < 0·0001; Years 2: 0·01; Year 3-5: >0.05; Dementia status: Years 1-3: < 0·0001, Year 4-5: 0·001 ; Diabetes status: Year 1: 0·0003; Years 2-5: < 0·0001; Pathogen of severe infection exposure: Years 1-5: > 0·05; Type of severe infection exposure: Years 1-5: < 0·0001.  CI = confidence interval, eFI = Electronic Frailty Index. | | | | | | | | | | |

| **Table S9. England cohort: Prevalence of Electronic Frailty Index deficits over time** | | | | | | | | | | | | |
| --- | --- | --- | --- | --- | --- | --- | --- | --- | --- | --- | --- | --- |
|  | **Index date** | | **Year 1** | | **Year 2** | | **Year 3** | | **Year 4** | | **Year 5** | |
|  | **No severe infection at index date** | **Severe infection at index date** | **No severe infection at index date** | **Severe infection at index date** | **No severe infection at index date** | **Severe infection at index date** | **No severe infection at index date** | **Severe infection at index date** | **No severe infection at index date** | **Severe infection at index date** | **No severe infection at index date** | **Severe infection at index date** |
|  | n = 1,208,3741 | n = 243,7431 | n = 1,208,3741 | n = 243,7431 | n = 1,031,1131 | n = 160,8481 | n = 869,3541 | n = 125,8731 | n = 725,0161 | n = 99,0591 | n = 600,8111 | n = 77,8171 |
| Activity limitation | 21,865 (1·8%) | 9,269 (3·8%) | 25,648 (2·1%) | 11,588 (4·8%) | 22,578 (2·2%) | 7,043 (4·4%) | 19,672 (2·3%) | 5,521 (4·4%) | 16,870 (2·3%) | 4,362 (4·4%) | 14,278 (2·4%) | 3,503 (4·5%) |
| Anaemia | 148,551 (12%) | 48,231 (20%) | 167,553 (14%) | 55,659 (23%) | 150,331 (15%) | 37,996 (24%) | 132,881 (15%) | 30,851 (25%) | 115,873 (16%) | 25,024 (25%) | 100,236 (17%) | 20,039 (26%) |
| Arthritis | 451,757 (37%) | 106,286 (44%) | 472,033 (39%) | 109,444 (45%) | 416,318 (40%) | 75,632 (47%) | 362,033 (42%) | 61,098 (49%) | 310,789 (43%) | 49,546 (50%) | 264,942 (44%) | 40,039 (51%) |
| Atrial fibrillation | 110,176 (9·1%) | 35,625 (15%) | 124,058 (10%) | 41,498 (17%) | 110,769 (11%) | 27,431 (17%) | 97,190 (11%) | 21,779 (17%) | 84,820 (12%) | 17,412 (18%) | 73,598 (12%) | 13,908 (18%) |
| Cerebrovascular disease | 109,111 (9·0%) | 34,706 (14%) | 119,644 (9·9%) | 38,251 (16%) | 103,500 (10%) | 25,436 (16%) | 88,656 (10%) | 19,930 (16%) | 75,528 (10%) | 15,729 (16%) | 64,257 (11%) | 12,403 (16%) |
| Chronic kidney disease | 209,085 (17%) | 57,348 (24%) | 240,140 (20%) | 64,263 (26%) | 218,374 (21%) | 44,474 (28%) | 193,851 (22%) | 36,349 (29%) | 168,912 (23%) | 29,558 (30%) | 145,602 (24%) | 23,783 (31%) |
| Dizziness | 223,437 (18%) | 51,813 (21%) | 239,829 (20%) | 54,824 (22%) | 216,828 (21%) | 38,638 (24%) | 192,303 (22%) | 31,718 (25%) | 168,244 (23%) | 26,000 (26%) | 145,641 (24%) | 21,236 (27%) |
| Dyspnoea | 226,481 (19%) | 80,733 (33%) | 252,990 (21%) | 88,333 (36%) | 229,335 (22%) | 61,798 (38%) | 204,305 (24%) | 50,083 (40%) | 178,978 (25%) | 40,514 (41%) | 154,626 (26%) | 32,286 (41%) |
| Falls | 171,219 (14%) | 53,901 (22%) | 198,905 (16%) | 63,142 (26%) | 178,872 (17%) | 42,488 (26%) | 158,597 (18%) | 34,209 (27%) | 139,518 (19%) | 27,689 (28%) | 121,784 (20%) | 22,227 (29%) |
| Foot problems | 99,204 (8·2%) | 31,545 (13%) | 109,889 (9·1%) | 34,830 (14%) | 99,538 (9·7%) | 24,483 (15%) | 89,089 (10%) | 20,154 (16%) | 78,436 (11%) | 16,566 (17%) | 68,607 (11%) | 13,514 (17%) |
| Fragility fracture | 141,702 (12%) | 37,962 (16%) | 154,370 (13%) | 42,052 (17%) | 134,980 (13%) | 27,944 (17%) | 116,515 (13%) | 22,240 (18%) | 99,663 (14%) | 17,690 (18%) | 85,295 (14%) | 14,072 (18%) |
| Hearing loss | 197,715 (16%) | 42,833 (18%) | 213,704 (18%) | 45,874 (19%) | 191,915 (19%) | 31,693 (20%) | 169,755 (20%) | 25,980 (21%) | 148,154 (20%) | 21,332 (22%) | 128,594 (21%) | 17,490 (22%) |
| Heart failure | 64,337 (5·3%) | 25,395 (10%) | 74,177 (6·1%) | 30,563 (13%) | 65,285 (6·3%) | 19,931 (12%) | 56,993 (6·6%) | 15,790 (13%) | 49,253 (6·8%) | 12,614 (13%) | 42,726 (7·1%) | 9,955 (13%) |
| Heart valve disease | 17,284 (1·4%) | 5,583 (2·3%) | 20,054 (1·7%) | 6,448 (2·6%) | 18,363 (1·8%) | 4,320 (2·7%) | 16,669 (1·9%) | 3,499 (2·8%) | 14,745 (2·0%) | 2,871 (2·9%) | 13,130 (2·2%) | 2,389 (3·1%) |
| Housebound | 158,792 (13%) | 60,799 (25%) | 188,323 (16%) | 76,308 (31%) | 165,300 (16%) | 50,222 (31%) | 143,941 (17%) | 40,183 (32%) | 124,399 (17%) | 32,159 (32%) | 106,703 (18%) | 25,311 (33%) |
| Hypertension | 605,997 (50%) | 131,823 (54%) | 623,123 (52%) | 134,273 (55%) | 544,842 (53%) | 90,040 (56%) | 469,642 (54%) | 71,619 (57%) | 399,963 (55%) | 57,254 (58%) | 337,989 (56%) | 45,626 (59%) |
| Hypotension syncope | 132,871 (11%) | 38,523 (16%) | 148,121 (12%) | 43,691 (18%) | 133,238 (13%) | 30,071 (19%) | 118,215 (14%) | 24,467 (19%) | 103,532 (14%) | 19,881 (20%) | 89,812 (15%) | 16,105 (21%) |
| Ischemic heart disease | 162,487 (13%) | 43,888 (18%) | 167,859 (14%) | 45,280 (19%) | 145,479 (14%) | 30,540 (19%) | 124,902 (14%) | 24,348 (19%) | 105,944 (15%) | 19,580 (20%) | 89,352 (15%) | 15,624 (20%) |
| Memory cognitive problems | 119,197 (9·9%) | 35,756 (15%) | 144,825 (12%) | 46,086 (19%) | 125,646 (12%) | 30,116 (19%) | 108,816 (13%) | 23,882 (19%) | 94,603 (13%) | 19,124 (19%) | 82,942 (14%) | 15,435 (20%) |
| Mobility transfer problems | 82,903 (6·9%) | 33,495 (14%) | 95,466 (7·9%) | 39,110 (16%) | 83,812 (8·1%) | 27,051 (17%) | 72,878 (8·4%) | 22,025 (17%) | 62,519 (8·6%) | 17,770 (18%) | 53,499 (8·9%) | 14,194 (18%) |
| Osteoporosis | 97,325 (8·1%) | 25,843 (11%) | 105,741 (8·8%) | 28,258 (12%) | 93,854 (9·1%) | 19,811 (12%) | 82,302 (9·5%) | 16,161 (13%) | 71,359 (9·8%) | 13,024 (13%) | 61,501 (10%) | 10,474 (13%) |
| Parkinson’s tremors | 36,131 (3·0%) | 12,770 (5·2%) | 40,115 (3·3%) | 14,217 (5·8%) | 35,234 (3·4%) | 9,596 (6·0%) | 30,627 (3·5%) | 7,663 (6·1%) | 26,444 (3·6%) | 6,009 (6·1%) | 22,615 (3·8%) | 4,767 (6·1%) |
| Peptic ulcer | 55,538 (4·6%) | 15,733 (6·5%) | 57,458 (4·8%) | 16,810 (6·9%) | 49,573 (4·8%) | 11,193 (7·0%) | 42,366 (4·9%) | 8,939 (7·1%) | 36,031 (5·0%) | 7,076 (7·1%) | 30,260 (5·0%) | 5,601 (7·2%) |
| Peripheral vascular disease | 40,028 (3·3%) | 14,888 (6·1%) | 43,117 (3·6%) | 16,063 (6·6%) | 37,868 (3·7%) | 10,671 (6·6%) | 32,803 (3·8%) | 8,501 (6·8%) | 28,009 (3·9%) | 6,796 (6·9%) | 23,716 (3·9%) | 5,363 (6·9%) |
| Polypharmacy | 657,188 (54%) | 156,974 (64%) | 762,778 (63%) | 183,771 (75%) | 702,922 (68%) | 120,452 (75%) | 642,602 (74%) | 99,696 (79%) | 590,373 (81%) | 84,998 (86%) | 570,884 (95%) | 76,891 (99%) |
| Requirement for care | 88,489 (7·3%) | 25,029 (10%) | 105,919 (8·8%) | 32,452 (13%) | 90,077 (8·7%) | 21,629 (13%) | 76,338 (8·8%) | 17,741 (14%) | 64,742 (8·9%) | 14,396 (15%) | 55,457 (9·2%) | 11,467 (15%) |
| Respiratory disease | 102,708 (8·5%) | 50,487 (21%) | 112,469 (9·3%) | 55,277 (23%) | 99,311 (9·6%) | 38,176 (24%) | 86,509 (10·0%) | 30,172 (24%) | 74,485 (10%) | 23,882 (24%) | 63,421 (11%) | 18,681 (24%) |
| Rheumatoid diseases | 41,633 (3·4%) | 11,919 (4·9%) | 44,717 (3·7%) | 12,509 (5·1%) | 39,959 (3·9%) | 8,581 (5·3%) | 35,294 (4·1%) | 6,970 (5·5%) | 30,869 (4·3%) | 5,594 (5·6%) | 26,770 (4·5%) | 4,465 (5·7%) |
| Skin ulcer | 52,699 (4·4%) | 23,018 (9·4%) | 60,643 (5·0%) | 27,315 (11%) | 53,809 (5·2%) | 18,537 (12%) | 47,094 (5·4%) | 14,984 (12%) | 40,713 (5·6%) | 12,143 (12%) | 35,069 (5·8%) | 9,708 (12%) |
| Sleep disorders | 108,942 (9·0%) | 29,724 (12%) | 117,481 (9·7%) | 32,454 (13%) | 103,903 (10%) | 22,787 (14%) | 90,515 (10%) | 18,490 (15%) | 77,935 (11%) | 14,946 (15%) | 66,762 (11%) | 11,948 (15%) |
| Social vulnerability | 93,538 (7·7%) | 26,320 (11%) | 105,552 (8·7%) | 30,675 (13%) | 94,069 (9·1%) | 20,712 (13%) | 83,063 (9·6%) | 16,854 (13%) | 72,820 (10%) | 13,741 (14%) | 63,176 (11%) | 11,086 (14%) |
| Thyroid disease | 123,309 (10%) | 29,249 (12%) | 128,431 (11%) | 30,709 (13%) | 112,083 (11%) | 20,916 (13%) | 96,508 (11%) | 16,757 (13%) | 82,383 (11%) | 13,478 (14%) | 69,617 (12%) | 10,770 (14%) |
| Urinary incontinence | 77,477 (6·4%) | 24,832 (10%) | 86,033 (7·1%) | 28,235 (12%) | 75,457 (7·3%) | 19,985 (12%) | 65,340 (7·5%) | 16,238 (13%) | 56,022 (7·7%) | 13,141 (13%) | 47,714 (7·9%) | 10,602 (14%) |
| Urinary system disease | 301,297 (25%) | 79,359 (33%) | 321,926 (27%) | 86,914 (36%) | 287,412 (28%) | 62,113 (39%) | 252,522 (29%) | 50,715 (40%) | 218,870 (30%) | 41,617 (42%) | 188,248 (31%) | 33,906 (44%) |
| Visual impairment | 435,394 (36%) | 100,661 (41%) | 466,550 (39%) | 106,405 (44%) | 416,715 (40%) | 72,788 (45%) | 366,806 (42%) | 59,059 (47%) | 317,966 (44%) | 47,982 (48%) | 274,162 (46%) | 38,825 (50%) |
| Weight loss anorexia | 59,358 (4·9%) | 20,294 (8·3%) | 70,150 (5·8%) | 23,744 (9·7%) | 62,660 (6·1%) | 15,666 (9·7%) | 55,170 (6·3%) | 12,775 (10%) | 47,981 (6·6%) | 10,338 (10%) | 41,545 (6·9%) | 8,327 (11%) |
| Data are n (%). Percentages were calculated with the total number in the respective column headers (i.e., those at risk during follow-up) as denominators. | | | | | | | | | | | | |
